# Tracking antigen-specific T cell responses in patients with myocardial infarction

**DOI:** 10.1101/2025.06.05.25328514

**Authors:** Anna Rizakou, Michelle Bauer, Murilo Delgobo, Alexander Hofmann, Margarete Heinrichs, Johanna Siegel, Tobias Krammer, Antoine-Emmanuel Saliba, Kenz Le Gouge, Roland Jahns, Valerie Boivin-Jahns, Viacheslav Nikolaev, Ulrich Hofmann, Encarnita Mariotti-Ferrandiz, DiyaaElDin Ashour, Stefan Frantz, Gustavo Campos Ramos

**Affiliations:** Department of Internal Medicine I, University Hospital Würzburg, Würzburg, Germany; Comprehensive Heart Failure Center, University Hospital Würzburg, Würzburg, Germany; Helmholtz Institute for RNA-based Infection Research (HIRI), Helmholtz-Center for Infection Research (HZI), Würzburg, Germany; Institute of Molecular Infection Biology, Faculty of Medicine, University of Würzburg, Würzburg, Germany; Immunoregulation-Immunopathology-Immunotherapy, UMRS959, Sorbonne Université, INSERM, Paris, France; Interdisciplinary Bank of Biomaterials and Data Würzburg (IBDW), University Hospital Wurzburg, Würzburg, Germany; Institute of Pharmacology and Toxicology, University of Würzburg, Würzburg, Germany; Institute of Experimental Cardiovascular Research, University Medical Center Hamburg-Eppendorf, Hamburg, Germany; DZHK (German Center for Cardiovascular Research), Partner Site Hamburg/Kiel/Lübeck, Hamburg, Germany

**Keywords:** cardiac antigens, myocardial infarction, TCR, adrenergic receptor, T cells, HLA-DRB1

## Abstract

Antigen-specific T cells can impact myocardial diseases, but appropriate model antigens and tools to track specific cells in patients are still lacking. Herein, we sought to identify and functionally characterize specific TCRs recognizing an epitope derived from the human adrenergic receptor beta 1 (ADRB1) presented on HLA-DRB1*13. Peripheral mononuclear cells obtained from HLA-DRB1*13^+^ subjects were stimulated in vitro with ADRB1_167-182_ peptide and the expanded CD4^+^ T cells expressing activation induced markers were sorted for single-cell RNA / TCR-sequencing. Downstream analysis revealed clonal expansion of ADRB1_167-182_-stimulated CD4^+^ T cells with a distinct TCR repertoire signature. Moreover, analysis of the TCRβ complementarity determining region 3 (CDR3) led to the identification of a distinct motif enriched in ADRB1_167-182_-stimulated IFN-γ-producing cells. Selected TCRs were expressed in reporter cell lines, and their antigen specificity was further assessed by specific HLA-tetramers and antigen stimulation. Moreover, immunophenotyping of peripheral blood cells from MI patients revealed that ADRB1-specific cells preferentially exhibited a memory phenotype. Taken together, our study provides annotation and functional validation of defined TCRs specific to an antigen relevant to human cardiovascular diseases, offering new tools to track antigen-specific cells in patients with myocardial disease.

## Main

Antigen-specific T cells impact the onset and progression of various myocardial diseases, including experimental autoimmune myocarditis ^1,2,3^, post-myocardial infarction (MI) repair ^4, 5, 6^ , pressure-overload-induced heart failure (HF) ^7,8^ and myocarditis induced by immune checkpoint inhibitors ^9,10^. These responses can either amplify tissue damage ^6, 11^ or orchestrate healing ^5^, requiring refined models to track and understand their complex time- and context-dependent functions. Myosin heavy chain alpha isoform (MYHCA) is a well-established cardiac antigen in defined mouse strains ^12,13^ and cardiac injury settings ^14, 9, 4^. However, the absence of model antigens and tools to study antigen-specific T cells in patients has impeded translational progress in cardio-immunology. Herein, we modelled T cell receptor (TCR) motifs specific for an epitope in the human adrenergic receptor beta 1 (ADRB1) presented on HLA-DRB1*13 and validated new tools to track specific T helper cells in myocardial disease patients.

The adult mammalian heart is characterized by low cell turnover and negligible regenerative capacity, and thus has limited tolerance to local inflammation ^15^. Consequently, tissue injury caused by MI – the leading cause of death worldwide – poses unique immunological challenges. In addition to the well-established roles of myeloid cells in mediating myocardial repair, T cells can also regulate cardiac inflammation in a complex, context-dependent manner ^16^. However, the absence of model cardiac antigens and defined tools to track heart-specific T cells in MI patients has hindered progress in translational cardio-immunology. In a previous study, we identified two overlapping peptide sequences derived from ADRB1 driving antigen-specific T cells responses in PBMC-derived CD4^+^ T cells from HLA-DRB1*13 patients with MI ^17^. Furthermore, ADRB1-specific antibodies have been described in different cohorts of myocardial disease patients ^18,19,20^.

With mounting evidence showing ADRB1 as a relevant autoantigen in MI and HF patients ^17,21^ and mice ^22,23^, we designed an experimental approach to expand and purify ADRB1_167-182_-specific CD4^+^ T cells to model specific TCR motifs, and thereby develop new tools to track specific T cells. Initially, PBMCs from HLA-DRB1*13^+^ subjects recruited in the frame of the “Cardiac antigens in MI” study ^17^ were chronically stimulated in vitro with ADRB1_167-182_ peptides to expand antigen-specific T cells for downstream purification. CD4^+^ T cells from HLA-DRB1*13^+^ subjects exhibiting the strongest response to ADRB1_167-182_-stimulation were then selected for single-cell RNA-/TCR-sequencing (Figure 1A). After initial screening, cells from five subjects (S1-S5) were selected based on surface expression of the AIM markers, OX40^+^ and 4-1BB^+ 24,25^ on T helper cells. Following antigen-driven clonal expansion, cells were multiplexed using 10 different hashtag antibodies and labelled with CITE-seq antibodies for CD25, CD69, CD45RA and CD45RO (Extended Data Figure 1). AIM^+^ T cells were sorted from both IL-2-control and ADRB1_167-182_-stimulated cultures and sequenced using the 5’ single-cell seq/TCR-seq 10x platform. After data filtering (Extended Data Figure 2A-B), 9,379 single CD4^+^ T cells were obtained, of which 8,315 showed a productive TCR sequence (Extended Data Figure 1B). Our analysis revealed 12 different clusters, including *naïve* T cells (expressing CD45^high^, *IL17R and CCR7*), 2 Treg clusters (expressing *FOXP3, IKZF2* and CD69^+/-^) and conventional T cells expressing effector/memory and cytotoxic genes (*KLRB1, GZMA* and *PTPN13*) (Figure 1B-C, Extended Data Figure 2C-D). Additionally, we identified a cluster associated with prototypical T_H_1 responses (expressing *IFNG*, *CCL3* and *GZMH*), a cluster with a T_H_2 signature (expressing *IL5, IL4* and *GATA3*), cells expressing the cytokines *CSF2* and *IL21* and a cluster of cells exhibiting high expression of *Il10* with checkpoint inhibitors and exhaustion genes (*LAG3, CTLA4, PDCD1* and *TOX*).

**Figure 1.**
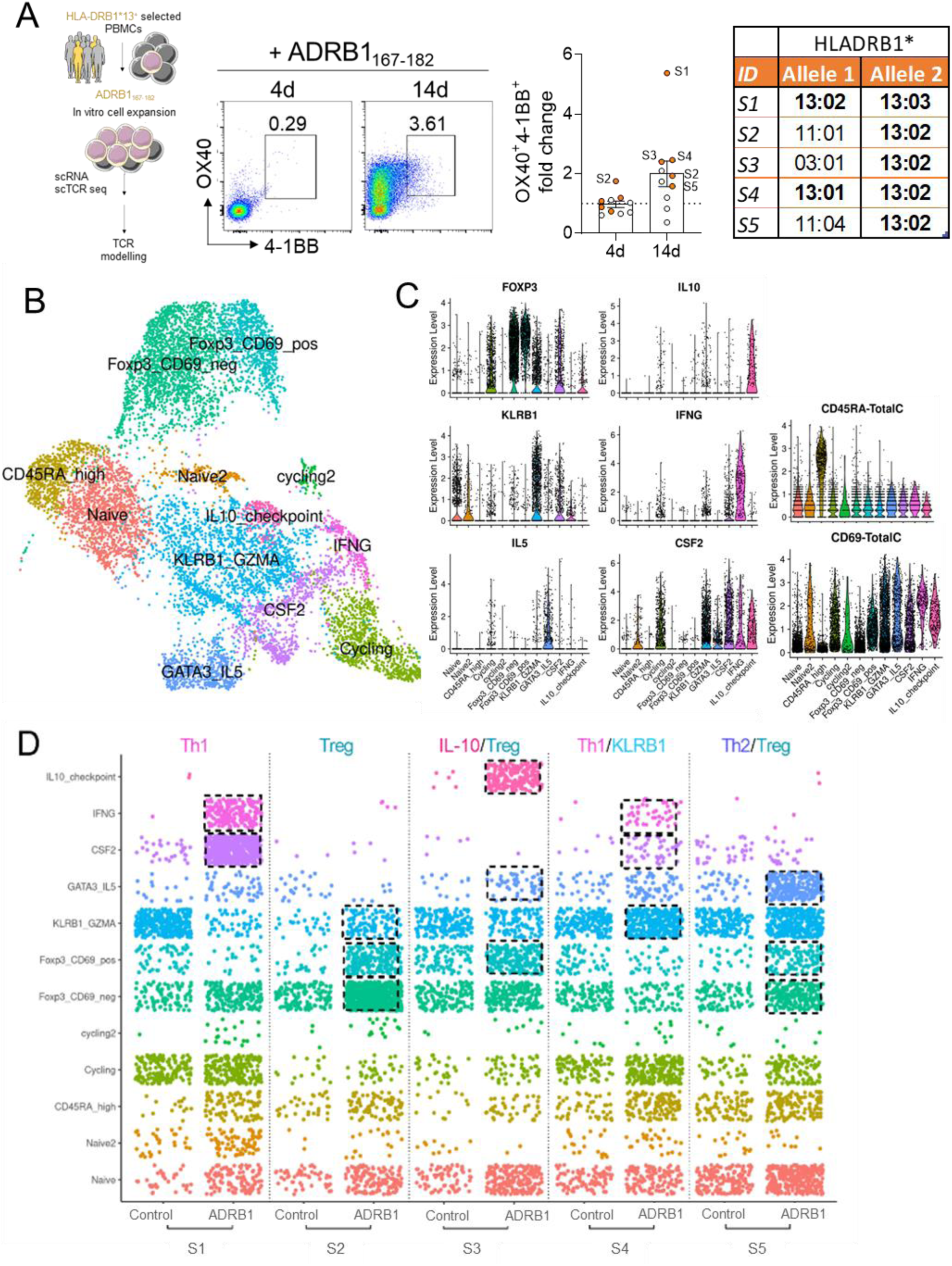
Single cell RNAseq profiling of human ADRB1 antigen-specific CD4^+^ T cell responses. (**A**) Experiment design: PBMCs of human subjects expressing the DRB1*13 HLA allele were stimulated in vitro with ADRB1_167-182_ peptide for 14 days. Donors showing the highest fold response in AIM T cells (OX40^+^4-1BB^+^) were selected for scRNA/ TCR seq analysis. The ‘dotted line highlight T cell subsets selectively expanded by the antigen. (**B**) UMAP plot displays 12 clusters annotated from CD4^+^ T cells derived from control and ADRB1-stimulated conditions. (**C**) Violin plots of the log-normalized expression values of *FOXP3, IL10, KLRB1, IFNG, IL5* and *CSF2* per cluster (left and central panel) and the CITE-seq signal for CD69 and CD45RA (right panel). (**D**) Jitter plots show cell distributions for each cluster defined in 1B, split by subjects (S1-S5) and experimental conditions (IL-2 control or ADRB1 peptide). Dashed squares highlight the most expanded clusters in ADRB1-stimulated condition for each subject.

Despite being maintained for two weeks in cell culture, the ADRB1_167-182_-expanded T helper cells showed a distinct transcriptome profile that was largely subject-specific. For instance, S1 and S4 had a skewed antigen specific T cell response enriched for T_H_1/KLRB1-associated transcripts. In contrast, S2 and S3 displayed a response dominated by Treg/IL-10 cells, whereas S5 exhibited clonally expanded cells with T_H_2/Treg response signatures (Figure 1D). These findings confirm that ADRB1_167-182_ is an HLA-DRB1*13-restricted autoantigen and that subject-specific responses can be tailored to a pro-inflammatory, regulatory or T_H_2 phenotype.

To investigate if the cluster-specific responses were associated with expansion of distinct TCRs, we evaluated clonal size distribution in control (IL-2 only) and peptide-stimulated groups. Our analysis revealed that more than 50% of TCR sequences were classified as medium to largely expanded in ADRB1_167-182_-stimulated cells, in sharp contrast to the TCRs found in controls (Figure 2A). We further confirmed these observed ADRB1_167-182_-induced clonal expansions by querying inverted Simpson’s index, which indicated a less diverse TCR repertoire in ADRB1_167-182_-stimulated cells (Extended Data Figure 3A). Overall, the TCR repertoire of peptide-stimulated cells can be broadly sorted into two distinct categories: a) antigen-specific responses dominated by a few highly expanded clones, mostly with a pro-inflammatory signature (e.g., in S1, S3 and S4); or b) antigen-specific responses comprised of medium-expanded clones, as typically observed in subjects with a Treg-biased response (e.g., S2 and S5) (Extended Data Figure 3B).

**Figure 2.**
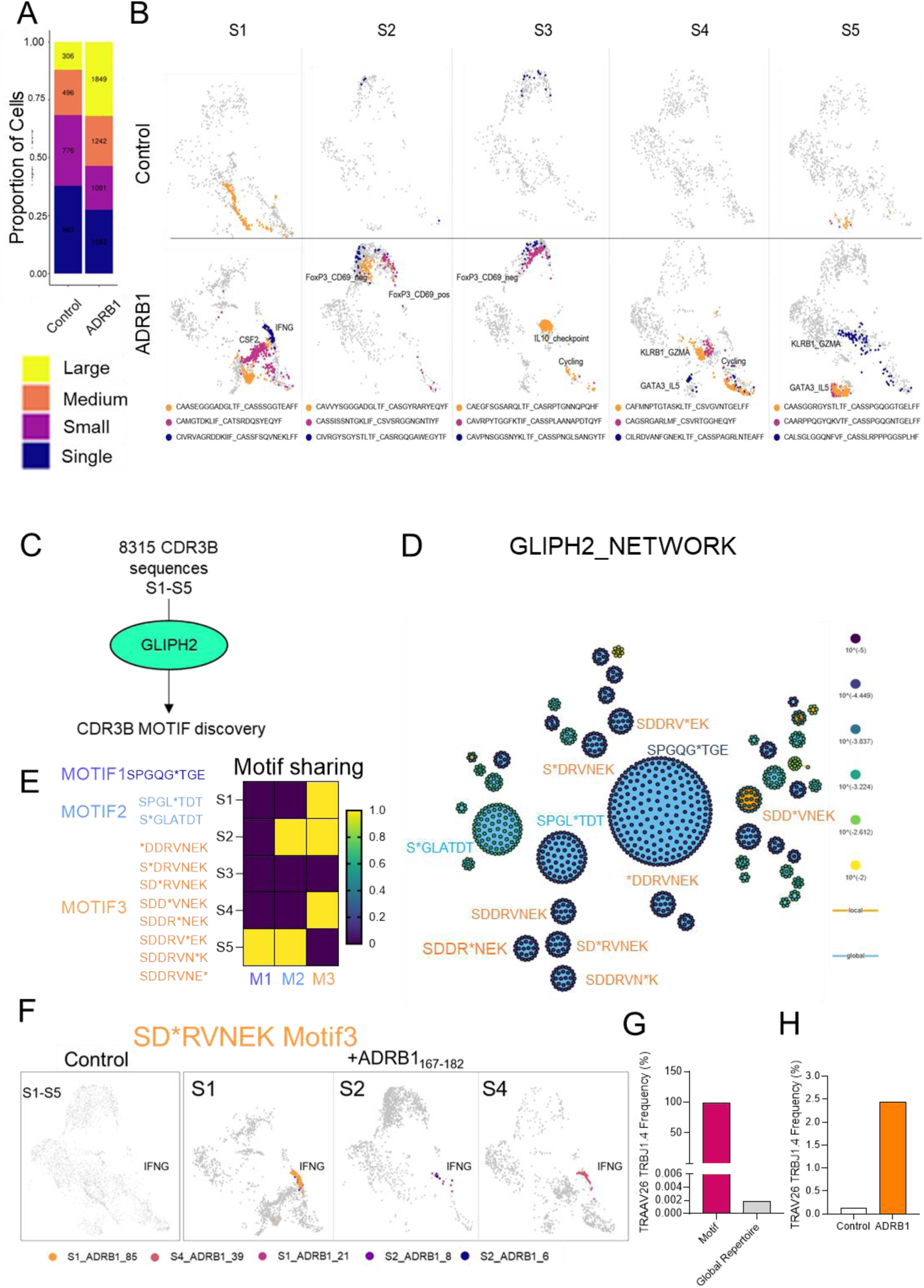
Clonal expansion, repertoire analysis of T cells and identification of CDR3β motifs in ADRB1 stimulated cells. (**A**) Graph shows the proportion of expanded clones in IL-2 control and ADRB1 peptide-stimulated groups (Large: 50<x≤400, Medium: 10<x≤50, Small: 1<x≤10, Single: 0<x≤1). (**B**) UMAP plots highlighting, per subject, the top three expanded clones. Sequences are in descending order; yellow is the top expanded and dark blue is the top three expanded. (**C**) Workflow of motif enrichment using GLIPH2 algorithm. (**D**) Network plot shows the most frequent (ball size) and with highest score (colour code) enriched motifs identified by GLIPH2. (**E**) Motifs identified by GLIPH2, and the respective subjects sharing the motif illustrated by heat map. (**F**) UMAP plots shows five TCR sequences sharing the motif 3 or the J1-4 sequence “NEKLFF”. Frequency of TRAAV26 TRBJ1.4 within the motif and global repertoire (**G**) and in control and ADRB1-stimulated cells (**H**).

Next, we aligned the top three expanded clones per subject to their respective cluster in the UMAP plot to identify potential ADRB1-specific TCRs. The top expanded TCR found in S1 was present in both control and ADRB1_167-182_-stimulation conditions, and its sequence was previously annotated to recognize a cytomegalovirus antigen ^26,27^. However, all other expanded TCR sequences identified in our dataset were exclusively found upon ADRB1_167-182_-stimulation, suggesting they might confer specificity to this antigen. In S1, the second and third most-expanded TCRs were enriched in ADRB1_167-182_-expanded T cells constituting the CSF2 and IFNG clusters (Figure 2B and 1D). Similarly, the top expanded clones found in S2 and S3 were mapped to Tregs/IL-10 clusters under the ADRB1_167-182_ stimulation regimen, whereas the top expanded clones found in S5 were mapped to T_H_2 cells (Figure 2B). A list of the top expanded clones in ADRB1-stimulated cells are shown in Table I and Extended Data Table I. Taken together, our data reveal that ADRB1_167-182_ peptide stimulation led to clonal expansions of CD4^+^ T cells in HLA-DRB1*13^+^ subjects.

**Table I.**
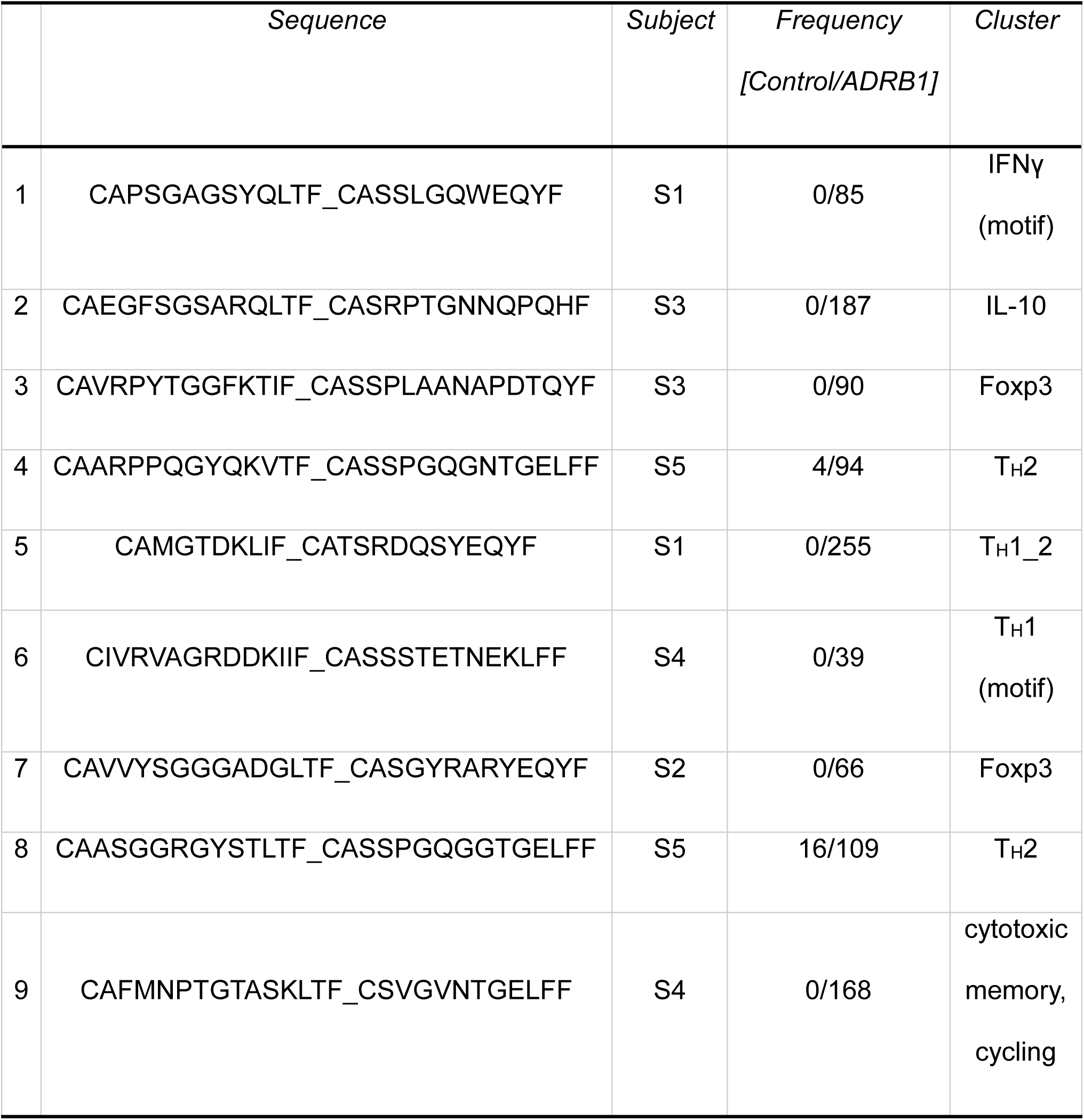
Top nine TCR α/β sequences expanded in ADRB1-stimulated PBMCs. For each sequence, the main representative cluster, subject and frequency are shown.

The TCR complementary-determining region 3 (CDR3) encompasses the antigen-binding sites and the rearrangement sites of TCR β variable (V) diversity (D) and joining (J) segments, and TCRα VJ segments, which account for most TCR variability. Applying a sequence-based metric, the Levenshtein distance which computes the number of amino acid substitutions, insertions or deletions between two given TCR CDR3β sequences, we estimated the sequence similarities between the top 50 CDR3β sequences found in ADRB1_167-182_-stimulated cells. As shown in Extended Data Figure 2E, while the top two expanded clones found in S5 exhibited great similarity, all remaining TCR sequences analyzed showed greater Levenshtein distances, thereby indicating that unique CDR3β TCR sequences largely characterize the ADRB1-specific TCR repertoire.

Because the clonally expanded TCRs were mostly subject-specific private, we next sought to identify TCR core motifs that may be shared among individuals (public), indicative of shared antigen recognition patterns. For this purpose, 8,315 CDR3β sequences derived from control and peptide-stimulated cells were analyzed using the GLIPH2 algorithm (Figure 2C), which identifies conserved sequences enriched in datasets of interest against a reference repertoire of 162,165 human TCRs and represented in the network using a composite “total_score” parameter. The total_score integrates motif enrichment, HLA association, and inter-individual prevalence ^28,29^. The top two most frequent CDR3β motifs (SPGQG*TGE; SPGL*TDT) were mainly shared within S5 and show no common CDR3α gene usage. However, the third most frequent CDR3β motif (SD*RVNEK, motif 3) showed the highest Fischer score (7.5e-08) and several additional related variants (e.g., S*DRVNEK, SDDRV*EK) were also identified (Figure 2D-E). Building on these observations, we interrogated the distribution and phenotype of T cells expressing this CDR3β motif. Strikingly, motif 3 was present only in ADRB1_167-182_-stimulated cell cultures and was shared between sequences from S1 and S2 subjects (Figure 2F). We then expanded our investigation to cells expressing “NEKLFF”, the germline sequence encoded by TRBJ1-4. With these new criteria, we found two additional sequences from S1 and S4 that were 85 and 39 times expanded, respectively (Figure 2F). All T cells expressing the CDR3β motif 3 also shared the exact same CDR3α TRAV26-1_TRAJ30 segment usage (Figure 2F-G), further confirming that this might be a bona fide ADRB1-specific TCR motif. Intriguingly, all T cells expressing this distinct TCR motif aligned within the IFNG cluster, even in S2, that showed a dominant Treg response (Figure 2F, 1D and 2B). Moreover, the TRAV26-1_TCBJ1-4 gene segment usage was enriched among ADRB1-stimulated cells, in sharp contrast with the global TCR repertoire (Figure 2H), suggesting this particular TCR αβ combination might be enriched for specificity against the HLA-DRB1*13:ADRB1_167-182_ epitope. Taken together, these results demonstrate that motif 3 identifies TCR sequences conserved amongst individuals and exclusively found in ADRB1-stimlated T cells posed with a T_H_1 signature.

Next, to validate this identified TCR signature (Extended Data Table I), we selected nine TCRs (Table I) to be expressed on a modified Thymoma cell line (BW5147. G.1.4) and tested in vitro ^30^. In brief, to avoid mismatching between the endogenous and introduced TCRs, we employed CRISPR/Cas9 mediated knockout of the endogenous TCRα constant chain. Then we retrovirally introduced the complete CD3 complex (CD3γ, CD3δ, CD3ε, and CD3ζ) ^31^ and human mutated CD4 (hmuCD4) ^32^ in order to generate a suitable cell line (BW.CD3.CD4.NFAT-ZsGreen) to express and test the transgenic TCRs identified in our scTCR datasets (Figure 3A). TCR activation was assessed by ZsGreen Signal reporter under the NFAT promoter.

**Figure 3:**
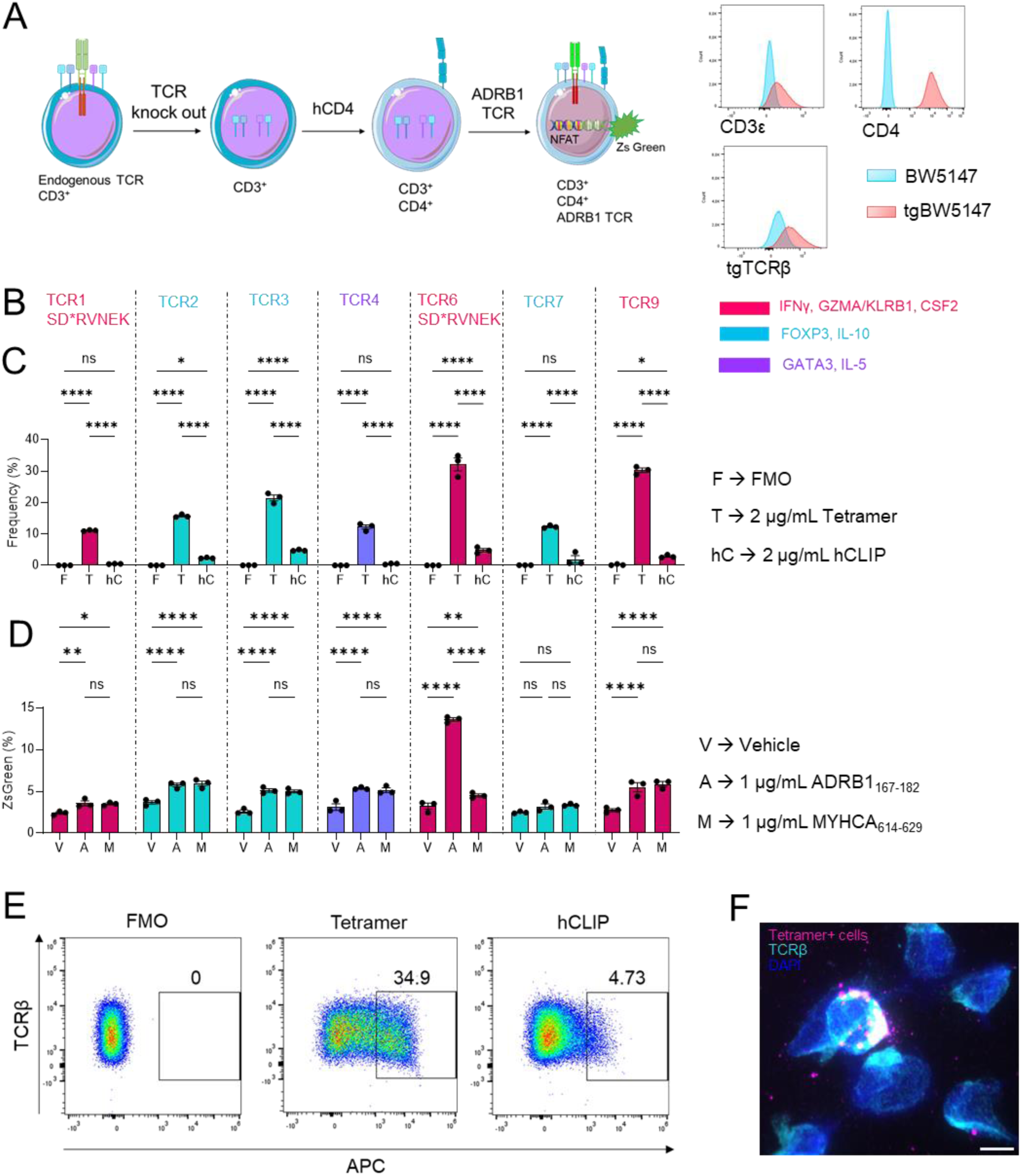
Functional validation of predicted TCR motifs. (**A**) Workflow of tgTCR cell line generation (left panel) and representative flow cytometry histograms of unmodified BWs (cyan) versus tgTCR BWs (red) (right panel). (**B**) The seven tetramer-enriched generated tgTCRs, coloured according to their corresponding cell cluster from scRNAseq (Figure 2B). TCR1 and TCR6 express the SD*RVNEK motif described in Figure 2. (**C**) Staining of tetramer-enriched tgTCRs with MHCII-restricted tetramer against ADRB1_167-182_ and control hCLIP peptide. Unstained tgTCRs served as FMO. (n=3 technical replicates; data are shown as mean±SEM; statistical analysis was performed with two-way ANOVA followed by Tukey’s multiple comparison test. *P*-values < 0.05 were considered statistically significant with: ***, *P*<0.001; ****, *P*<0.0001). (**D**) Co-culture of ADRB1_167-182_ specific tgTCRs with PBMCs from matching donor (ratio 1:1) in presence of 1 µg/mL ADRB1_167-182_, 1 µg/mL MYHCA_614-629_ or vehicle. (n=3 technical replicates; data are shown as mean±SEM; statistical analysis was performed with two-way ANOVA followed by Tukey’s multiple comparison test. *P*-values < 0.05 were considered statistically significant with: ****, *P*<0.0001). (**E**) Representative flow cytometry plots of tetramer-enriched TCR6 unstained (left), stained with tetramer (center) or hCLIP (right). (**F**) Representative fluorescent image of TCR6 cells stained by immunofluorescence for tetramer+ (magenta), TCRβ (cyan) and DAPI (blue). Images were acquired on a Leica Imager DMi8 equipped with a 63x objective. Scale bar: 10 μm.

The nine selected TCRs of interest (Table I, Extended Data Figure 5B) were then retrovirally expressed in the in-house-generated cells. We obtained custom-made fluorescently labelled HLA-DRB1*13:02 MHCII tetramers loaded with ADRB1_167-182_, which allowed us to detect antigen-specific CD4^+^ T cells in samples from myocardial disease patients with matching HLA (kindly provided by the NIH Tetramer Core Facility). HLA-DRB1*13:02 tetramers loaded with the CLIP peptide were used as control. After establishing optimal tetramer staining conditions (Extended Data Figure 4), we assessed the specific binding of all nine TCRs clonotypes to HLA-DRB1*13:02:ADRB1_167-182_ tetramers. As shown in Extended Data Figure 5C, all tested TCR clones showed HLA-DRB1*13:02:ADRB1_167-182_ tetramer^+^ staining. TCR9 displayed the highest frequency of specific stained cells, followed by TCR4 and TCR6. The seven clonotypes exhibiting tetramer^+^ stained cell frequencies greater than 8% were selected for enrichment via fluorescence-activated cell sorting (FACS) (Extended Data Figure 5D) and the experiment was subsequently repeated using the tetramer-enriched clonotypes (Figure 3B-C and E). Notably, TCR6 and TCR9 again demonstrated the highest frequencies of HLA-DRB1*13:02:ADRB1_167-182_ tetramer^+^ cells. Interestingly, TCR9 and TCR6 originated from the same donor and were enriched in ADRB1-stimulated cells showing a pro-inflammatory phenotype (Figure 3B, Table I). The specificity of the tetramer staining was further confirmed by immunofluorescence imaging, which indicated colocalization of specific tetramer and TCRβ signal per cell base (Figure 3F, Extended Data Figure 6). To further validate the expressed TCRs, the tgTCR-BW.CD3.CD4.NFAT cells were co-cultured with matching HLA-DRB1*13^+^ PBMCs at a 1:1 ratio and stimulated with 1 μg/mL ADRB1_167-182_ or vehicle. A peptide derived from MYHCA isoform MYHCA_614-629_, irrelevant in this condition, was used as control (Extended Data Figure 7). As shown in Figure 3D, cells expressing TCR6 exhibited a specific response to ADRB1 stimulation, as evidenced by ZsGreen expression. Interestingly, this TCR6 clonotype comprises the conserved CDR3β motif 3 identified in Figure 2 (Figure 3B, Table I). The remaining clonotypes tested did not show significant ZsGreen signal upon ADRB1-stimulation (Figure 3D).

After identifying TCRs specific for an HLA-DRB1*13-restricted epitope enriched in the heart and validating distinct MHC-II tetramers to track specific cells, we monitored HLA-DRB1*13 frequencies in different cohorts of MI patients recruited in Würzburg ^33,17^ and compared them with available databases of German bone marrow donors ^34^. As shown in Figure 4A, the combined frequency of HLA-DRB1*13:01, HLA-DRB1*13:02 and HLA-DRB1*13:03 alleles was higher in MI patients (n=87) compared to the healthy German population (n= 3,456,066), though not statistically significant. Likewise, the fraction of individuals carrying these alleles was also enriched among MI patients (Figure 4B).

**Figure 4:**
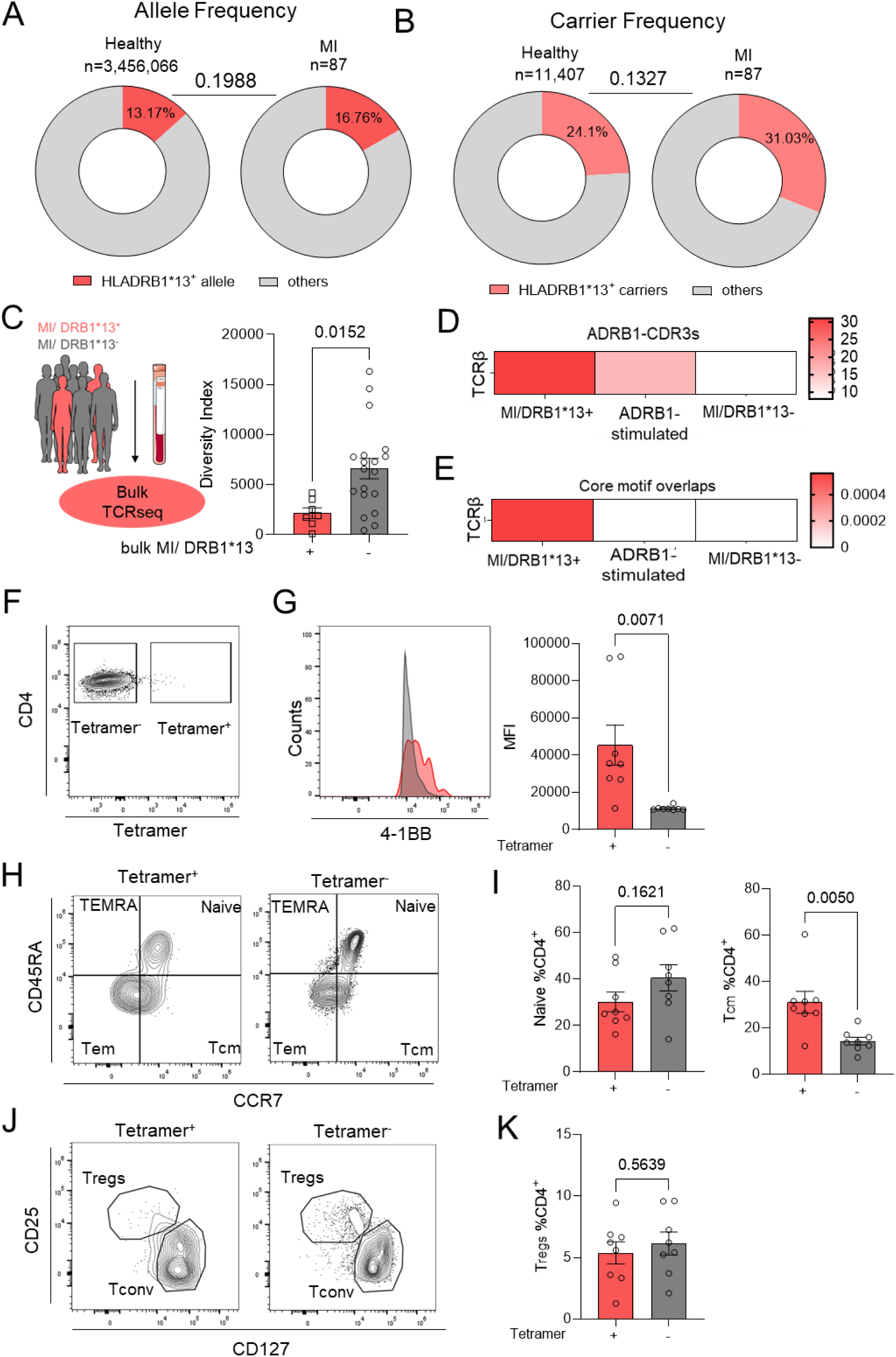
HLADRB1*13 allele as a potential risk factor for ADRB1 specific T cell responses post MI. (**A**) Donut charts depicting HLA-DRB1*13^+^ allele frequency in healthy population and MI patients. Statistical analysis was performed with Chi-square test followed by Yates’ correction (*P*=0.1988). *P*-values < 0.05 were considered statistically significant. (**B**) Donut charts depicting HLADRB1*13^+^ carrier frequency in healthy population and MI patients. Statistical analysis was performed with Fisher’s exact test (P= 0.1327). *P*-values < 0.05 were considered statistically significant. (**C**) Experimental workflow (left panel) and diversity index of TCRβ chain from the bulkRNAseq data of DRB1*13^+^ (n=7) versus DRB1*13^−^ (n=19) MI patients. Both groups displayed similar Troponin levels. Data are shown as mean±SEM; statistical analysis was performed with unpaired t-test. *P*-values < 0.05 were considered statistically significant. (**D**) Heatmap depicting the enrichment of TCRβ sequences from Table I in the CDR3s of MI/ DRB1*13^+^, ADRB1 stimulated PBMCs (from Hapke *et al.*) and MI/ DRB1*13^−^ bulkRNAseq data, without selection for expansion. (**E**) Heatmap depicting the core motif overlapping (>50 clones, ≥1e-04) within TCRβ sequences from Table I and MI/ DRB1*13^+^, ADRB1 stimulated PBMCs and MI/ DRB1*13^−^. (**F**) Representative flow cytometry plot of CD4/Tetramer gated on live CD45^+^/CD3^+^/CD4^+^ cells (left), (**G**) Histogram (right panel) and quantitative analysis of 4-1BB^+^ cells in tetramer^+^ versus tetramer^−^ cells for patient (n=8) (left panel). Data are shown as mean ± SEM; Statistical analysis was performed with unpaired t-test. *P*-values < 0.05 were considered statistically significant. (**H**) Representative flow cytometry plot of CD45RA/CCR7 gated on live CD45^+^/CD3^+^/CD4^+^ cells and (**I**) quantitative analysis of Naïve and central-memory T cells (T_cm_) in tetramer^+^ versus tetramer^−^ cells for each patient (n=8). Data are shown as mean ± SEM; Statistical analysis was performed with unpaired t-test. *P*-values < 0.05 were considered statistically significant. (**J**) Representative flow cytometry plot of CD25/CD127 gated on live CD45^+^/CD3^+^/CD4^+^ cells (left panel) and (**K**) quantitative analysis of Tregs in tetramer^+^ versus tetramer^−^ cells for each patient (n=8). Data are shown as mean ± SEM; Statistical analysis was performed with unpaired t-test. *P*-values < 0.05 were considered statistically significant.

To further characterize post-MI antigen-specific T cell responses in patients, we retrospectively analysed TCRseq data available from peripheral blood samples obtained from MI patients stratified by HLA genotype ^35,33^. TCR repertoire analysis performed on samples obtained at index hospitalization revealed reduced TCRβ diversity in DRB1*13^+^ MI patients, suggestive of antigen-driven clonal expansions in MI patients with this HLA genotype (Figure 4C, Extended Data Figure 8A). A similar pattern was also observed for TCRα sequences (Extended data Figure 8B-C). Moreover, we found that the TCR repertoires of DRB1*13^+^ MI patients were enriched for CDR3β sequences annotated to be ADRB1-specific, based on our TCR modelling (Figure 4D) and core motif overlaps (Figure 4E).

Using the HLA-DRB1*13:02 : ADRB1_167-182_ tetramer validated in Figure 3, we were able to track and immunophenotype antigen-specific T helper cells obtained from matching MI patients (Figure 4F-K, Extended Data Figure 9-10). Interestingly, tetramer^+^ CD4^+^ cells displayed higher surface expression levels of AIM marker 4-1BB^+^ (Figure 4F-G) and increased frequency of central memory T helper (CD3^+^CD4^+^CCR7^hi^CD45RA^lo^) cells (Figure 4H-I), compared to patient-matched tetramer^−^ cells. No significant differences in Treg frequency were observed between tetramer^+^ and tetramer^−^ cell populations (Figure 4J-K). These findings suggest that CD4^+^ T cells might be already primed against ADRB1-derived epitopes before MI onset, possibly in the context of atherosclerosis and coronary heart disease. This would explain the presence of central memory T cells in the early stages of MI, validating the higher frequency of HLA-DRB1*13 alleles and reduced TCR clonal diversity among MI patients. These results reinforce the need to understand heart-directed immune responses across the full ischemic heart disease spectrum, establishing ADRB1 as an autoantigen in coronary heart disease and MI.

The previous identification of APOB1-specific CD4^+^T cells and the development of specific MHC-II tetramers were game-changing advancements that enabled in-depth characterization of antigen-specific T cells in atherosclerosis ^36, 37, 38^. However, the lack of identified cardiac antigens in humans has limited our understanding of heart-directed immune responses in patients. Our study addresses this gap by functionally validating TCR motifs specific for an antigen relevant in patients with myocardial disease and a defined HLA composition. To the best of our knowledge, this is the first annotation of cardiac-specific TCRs validated in humans. A growing number of clinical studies have characterized T cells in MI patients, ^40^ and the TCR resources generated in our study will allow antigen specificity to be inferred from TCR-sequencing data, both retrospectively and prospectively. Additionally, the tetramers herein described will enable future diagnostic, prognostic and mechanistic interrogation of antigen-specific T cells in heart disease patients representing a significant progress in the field of translational cardio-immunology.

## Supporting information

Extended Data Table III

## Data Availability

The anonymised patient data can be shared under reasonable request. The RNA sequencing data will be deposited on a public repository after the peer review process is completed.

## List of non-standard abbreviations

ADRB1: adrenergic receptor beta 1
AIM: activation induced marker
CD: cluster of differentiation
CDR3: complementarity-determining region 3
CITE-seq: cellular indexing of transcriptomes and epitopes by sequencing
FACS: fluorescent activated cell sorting
FOXP3: forkhead box P3
GLIPH: grouping of lymphocyte interactions by paratope hotpots
HF: heart failure
HLA: human leukocyte antigen
IFN-γ: interferon gamma
IL: interleukin
MHC: major histocompatibility complex
MI: myocardial infarction
MYHCA: myosin heavy chain alpha isoform
PBMCs: peripheral blood mononuclear cells
scRNA-seq: single-cell RNA sequencing
scTCR-seq: single-cell T cell receptor sequencing
TCR: T cell receptor
T_cm_: T central memory cell
T_em_: T effector memory cell
T_EMRA_: effector memory cells re-expressing CD45RA
T_H_: T helper cell
TRA: T cell receptor alpha genes
TRB: T cell receptor beta genes,
Treg: regulatory T cell
V(D)J: variable diversity and joining gene segment.

## Online content

A detailed methodological description along with extended Figures and Tables are presented as online content.

## Methods

### Patient Cohorts

In the present study, we analysed cryopreserved samples available from three clinical studies conducted in Würzburg ^33,17^. First, we performed expansion of antigen-specific T helper cells and TCR characterization with peripheral blood mononuclear cells (PBMCs) from HLA-DRB1*13 patients previously recruited within the frame of the ‘Cardiac Antigens after MI’ (CAMI) study ^17^. CAMI included patients diagnosed with first acute MI, control patients with cardiovascular risk factors but without signs of coronary artery disease who underwent elective coronary angiography (CA group) and heathy subjects (HS group). In total, PBMCs from five HLA-DRB1*13^+^ subjects from the CAMI cohort were selected for scRNA/TCRseq experiments. In a second series of experiments, PBMCs from 8 MI HLA-DRB1*13^+^ subjects were used for spectral flow cytometry investigation. We tracked ADRB1-specific T cells and immunophenotyped the tetramer^+^ and tetramer^−^ CD4^+^ populations in 6 patients from the CAMI cohort and 2 patients from the ANTIK-MI („Antikörper-Produktion durch B-Zellen nach Myokardinfarkt”) study. Finally, we retrospectively analysed a TCR sequencing dataset available from our previous study. The Etiology, Titre-Course and Survival (ETiCS) cohort included 31 patients diagnosed with first acute MI, also stratified by HLA-DRB1 genotype ^35^. The characteristics of these patients are described in Extended Data Table II.

### Study approval

The studies were approved by the ethics commission of the University of Würzburg (CAMI: 304/17-m3, ETiCS: 186/09, ANTIK-MI: 249/20-am) and met the criteria of the Declaration of Helsinki. All patients provided written informed consent.

### Expansion of human ADRB1_167-181_-specific CD4^+^ T cells

Available CAMI cohort ^17^ PBMCs expressing one or two copies of the HLA-DRB1*13 MHC-II haplotype were thawed and resuspended in complete RPMI media supplemented with 10% of human AB serum and 10 U/mL of rhIL-2 (Peprotech). Cells were either seeded at 2×10^6^ cells/mL in 48-well plate (day 0) and assigned to control (media + IL-2 only) or stimulated with 1 µg/mL of ADRB1_167-181_ and 1 µg/mL of ADRB1_168-182_ for 4 days at 37°C, 5% CO_2_. On day 4, cells were washed and resuspended in fresh complete RPMI media supplemented with IL-2. The washing steps were repeated on days 6, 8 and 11 and cells were always reseeded into the original plate (meaning they were in contact with autologous adherent cells throughout the experiment). On day 13, cells were re-stimulated with ADRB1_168-182_ peptides as previously indicated, in complete RPMI media supplemented with IL-2, and on day 14 we proceeded with the detection and purification of antigen-specific T cells based on their surface expression of activation-induced markers, as detailed below ^41, 24, 25^.

### Identification and purification of ADRB1_167-181_-specific CD4^+^ T cells

In order to identify antigen specific T cells based on their surface expression of activation-induced markers (AIM), less than 10^6^ cells were collected on days 4, 8 and 14 and processed for flow cytometry analysis. In brief, cells were washed once with PBS and stained with fixable viability dye (Zombie Aqua™ Fixable Viability Kit, BioLegend) for 15 minutes at room temperature. Next, cells were washed with FACS buffer (PBS containing 1% BSA, 0.1% Sodium Azide and 1 mM EDTA) and resuspended in 50 μL of antibody master mix, containing: anti-CD3 BV421 (clone UCHT1, 1:40, ThermoFisher), -CD4 FITC (clone SK3, 1:40, ThermoFisher), -4-1BB PE (CD137) (clone 4B4, 1:20, ThermoFisher) and - OX40 (CD134) APC (clone OX40, 1:50, Miltenyi), and Human TruStain FcX™ (1:200, BioLegend). Cells were kept for 30 minutes at 4°C and were then washed with FACS buffer. Finally, cells were resuspended in PBS and acquired using a Attune NxT FACS machine. T helper cells showing activation-induced activation markers (4-1BB^+^OX40^+^) on day 14 post-stimulation were defined as putative ADRB1_167-181_ -specific cells, as previously reported ^41, 24, 25^.

### Single cell RNAseq/TCR sequencing

After expanding the ADRB1_167-181_-specific CD4^+^ T cells in vitro over 14 days and selecting the ones showing response to ADRB1_167-181_ stimulation, cells were purified with fluorescence-activated cell sorting and then used for single cell RNA- / TCR sequencing. Cells were washed with sterile PBS, resuspended in ∼100µL and transferred to sterile U-bottom 96-well plates for staining. All buffers and centrifugation steps were conducted at 4°C. After centrifugation, cells were washed once with 200µL of PBS and then resuspended in 100µL of efluor 780 viability dye (1:1000, ThermoFisher). Cells were incubated for 15 minutes at room temperature, protected from light. Cells were then washed in sterile azide-free MACS buffer (0.5% BSA, 2mM EDTA in PBS). Afterwards, cells were resuspended in individual antibody master mix (staining volume 100µL), containing anti-CD3, -CD4, -4-1BB, -OX40, Human TruStain FcX. Immune cell populations were identified by sequencing for cellular indexing of transcriptomes and epitopes (CITE-seq), a technique that uses oligonucleotide-tagged antibodies to integrate cellular protein and transcriptome data into one single-cell readout ^42^. Therefore, the following totalseqC antibodies were added to the mix: CD69 TotalSeqC (clone FN50, 1:150), CD25 TotalseqC (clone BC96, 1:150), CD45RA TotalSeqC (clone HI100, 1:150) and CD45RO TotalSeqC (clone UCHL1, 1:150). Further, to enable downstream de-multiplexing of the scRNA-/ TCRseq data from pooled samples, the following TotalSeqC hashtag antibodies were used: CO251 (S1_control), CO252 (S1_adrb1), CO253 (S2_control), CO254 (S2_adrb1), CO255 (S3_control), CO256 (S3_adrb1), CO257 (S4_control), CO258 (S4_adrb1), CO259 (S5_control) and CO260 (S5_adrb1). All hashtag antibodies belonged to the clone LNH-94/2M2 and were used at 1:100 concentration. In brief, the cells were incubated in Ab master mix for 30 minutes at 4°C. Cells were then washed by adding 150uL of pre-chilled MACS buffer, centrifuged at 400 x *g* and 4°C for 3 minutes and resuspended in 200µL of MACS buffer. To sort antigen-activated T cells, 4-1BB^+^OX40^+^ events were sorted from live CD3^+^CD4^+^ T cells, with one collection tube used per donor. Collection tubes (1.5 mL biopur Eppendorf tubes) were pre-coated with 10% albumin solution (Miltenyi) for 24h and filled with 30µL of MACS buffer pre-sorting. 2/3 of sorted cells were derived from the ADRB1-stimulated condition and 1/3 from the control IL-2 condition. Finally, 20,000 cells/donor (peptide + control) were mixed in a new Eppendorf tube, (total cell number=100,000) and concentration was adjusted to 1,000 cells/µL.

### Single cell RNAseq and library construction

Single cell RNA-sequencing Chromium controller was used to partition single cells into nanolitre-scale Gel Bead-InEmulsion (GEMs) and Chromium Next GEM Single Cell 5’ v1.1 kits for reverse transcription, cDNA amplification and library construction (10x Genomics). A SimpliAmp Thermal cycler was used for amplification and incubation steps (Applied Biosystems). Libraries were quantified by a Qubit 3.0 fluorometer (ThermoFisher) and quality was checked using a 2100 Bioanalyser with high sensitivity DNA kit (Agilent) in paired-end mode to reach at least 60000 reads per single cell for gene expression and 6000 reads for the T cell receptor repertoire, hashtags and CITE-seqs. The Cell Ranger Software suite (version 7.0.1, 10x Genomics) was used for sequence alignment, barcode processing and sample demultiplexing.

### scRNA-/TCRseq data analysis, TCR modelling and motif discovery

The gene count matrix derived from Cell Ranger was analysed using Seurat package (version 5.0.1). The individual cell barcodes, ten different hashtag oligo sequences and the four CITE-seq antibody sequences were considered during sample de-multiplexing. In addition, cells yielding more than 50,000 unique molecular identifiers (UMI), with more than 5% of mitochondrial RNA and less than 200 features, were also excluded. During sample de-multiplexing using the HTOdemux function, 1,325 cells were considered doublets, 33 cells were considered negative and after data cleanup, 9,588 cells out of 11,200 cells remained. Cell transcriptome was log-normalized and the top 2000 most variable genes were obtained using Seurat’s default pipeline. The uniform manifold approximation and projection for dimension reduction (UMAP) was generated by the 20 principal component and a 0.5 resolution in dimensionality reduction. The analysis resulted in 14 different clusters and the function *FindallMarkers* in default settings was then used to estimate the differentially expressed genes in between clusters. The list of marker genes for each cluster can be found in Extended Data Table III. Two cell clusters identified as “monocytes” and “gd-Tcells” were subset-out of the main Seurat object, resulting in a “filtered” objected containing 9,379 cells.

The VDJ sequences for each T cell receptor were added to the Seurat object using the *combineExpression*() function from scRepertoire R package (1.11.0). Cells were classified either as: single clones (1), small-expanded clones (1-10), medium expanded clones (10-50) or largely expanded (50-400 clones. The function *occupiedRepertoire*() was used to show the distribution of clonotype size between control and peptide-stimulated conditions. The function *clonalDiversity()* was used to estimate the diversity index between all samples, with clonecall parameter set as “strict” (meaning CDR3a_CDR3b) and n.boots: 8315 (total number of cells with a productive TCR chain). The inverted Simpson’s index was plotted in GraphPad Prism (version 10.2.1). The top three expanded clones per subject were manually selected and are shown in figure 2B and 2F (motif) using the *highlightClonotypes()* function. GLIPH2 ^28^ was used for motif discovery, using the *turboGliph* package (version 0.99.2). A tab-separated table containing CDR3β, Vb, Jb, CDR3α, subject and frequency was loaded for the GLIPH2 pipeline. The *gliph_combined* function was used, with local_method: fisher, with clustering and scoring method coming from GLIPH2.0. The clones with the top score motifs were overlaid on the UMAP plots for analysing their phenotypic and patient-specific behaviour. This also included sequences sharing the same Jb gene 1-4 (NEKLFF) within the motif SD*RVNEK (Figure 2F).

### TCR repertoire analysis of ETiCS and CAMI cohorts

Repertoire analysis of the ETiCS bulk TCR seq data ^35^ and computation of overlaps with other TCR datasets were performed using the immunarch R package (version 1.0.0). Briefly, the MiXCR output files of bulk TCR seq ETiCS samples were imported into R using the repLoad() function. Samples were grouped according to their HLA-DRB1 haplotyping into DRB1*13^+^ or DRB1*13^−^. Distribution of clonotype abundances between DRB1*13^+^ or DRB1*13^−^ groups was computed using the repClonality() function. This method estimates the relative contribution of clones to the repertoire, stratifying them into four abundance-based categories: Small (<0.01%), Medium (0.01–0.1%), Large (0.1–1%), and Hyperexpanded (>1%). Overall repertoire diversity estimates were computed using the repDiversity() function based on the D50 index, which estimates the minimal number of unique clonotypes required to account for at least 50% of all sequencing reads in a given sample. Lower D50 values indicate greater clonal expansion, where fewer dominant clones occupy most of the repertoire. To evaluate TCR sharing between groups, CDR3β sequences from DRB1*13^+^ or DRB1*13^−^ groups were combined with TCRβ sequences from Table I, and ADRB1 stimulated PBMCs ^17^. The function repOverlap() was used to compute pairwise Jaccard similarity based on exact CDR3β matches between groups. To account for varying repertoire sizes between bulk and single-cell datasets, a scaling factor was applied to normalize all repertoire sizes to 1 million. To compute motif overlaps of the core CDR3 region between the TCRβ sequences of the current manuscript and the different datasets, we used a modified version of repOverlap(), which computes overlaps of truncated CDR3β sequences after removing the first 3 and last 3 amino acids representing the germline-encoded sequences of the V and J genes.

### Generation of a T cell reporter line and expression of TCRs identified in silico

BW5147 G.1.4 cells were purchased from ATTC (Manassas) and modified as previously described ^30^. In brief, CRISPR/CAS9 technology was used to knock out the endogenous TCRα constant chain. Afterwards, we used murine MSCV retroviral vectors for CD3-2A_pMI-LO (Addgene plasmid # 153418; http://n2t.net/addgene:153418; RRID: Addgene_153418) ^31^ to express all CD3 chains, and a humanized CD4 construct with a NFAT-Zs-Green reporter (8xNFAT-ZsG-muhCD4, Addgene plasmid #162745; https://www.addgene.org/162745/; RRID: Addgene_162745) ^32^. The aforementioned plasmids were purchased from Addgene and kindly deposited by Maki Nakayama. Finally, individual TCRs of interest (Table I) were synthesized and virally expressed in the BW-TCR-KO-mCD3^+^-muhCD4^+^-NFAT-ZsGreen reporter cell line (tgTCR).

### Characterization of cell lines expressing transgenic TCRs of interest

Next, cells expressing the TCRs identified in silico were characterized. First, cells were washed and then stained with efluor 780 viability dye (ThermoFisher) for 15 min at room temperature. Next, cells were washed with FACS Buffer and resuspended in 25μL of antibody master mix containing: TCRβ BV421 (clone H57-597, 1:100, BioLegend), hCD4 PE (clone A161A1, 1:100, BioLegend), CD3ε PercpCy5.5 (clone 145-2C11, 1:100, BioLegend), TruStain FcX™ (anti-mouse CD16/32, 1:200, BioLegend). Cells were kept for 15 minutes at 4°C and were then washed with FACS buffer. Finally, cells were resuspended in FACS Buffer and acquired in Attune NxT FACS machine. The tetramer and hCLIP were kindly provided by the NIH Tetramer Core Facility. Cells were washed and plated in a 96-well plate. After centrifugation, cells were resuspended in 50 μL of RPMI complete (1%Penicillin/ Streptomycin, 1% MEM-NEAA, 1% Glutamax, 1% Sodium Pyruvate, 0.1% β-Mercaptoethanol) with 2, 6 or 18 μg/mL of Tetramer or hCLIP and kept for 30, 90 or 180 minutes at 4 or 37°C. Cells were then washed with RPMI and proceeded for surface staining.

### Functional validation of expressed TCRs

Human PBMCs were thawed in RPMI complete medium in 10 cm petri dishes one day prior. tgTCR were co-cultured with HLA-matching PBMCs in a 1:1 ratio in U-bottom 96-well plates in the presence of 1 μg/mL ADRB1_167-182_, (QSLLTRARARGLVCTV, JPT Peptide Technologies), 1 μg/mL MYHCA_614-629_ (RSLKLMATLFSTYASADR, JPT Peptide Technologies), or vehicle for 24 hours. Cells were stained for viability, CD4, CD3ε and TCRβ and analysed by flow cytometry for NFAT-ZsGreen reporter activity.

### Validation of tetramer staining

In order to confirm tetramer specificity for ADRB1-specific T cells, we performed immunofluorescent staining to validate co-localization of tetramer (APC) with TCRβ. tgTCR cells from clone TCR6 were plated and left to rest overnight in an 8-well removable chamber plate (Ibidi) pre-coated with poly L-lysine (Sigma) to allow cell adherence, according to the manufacturer’s instructions. The following day, the medium was replaced with RPMI complete containing 2 μg/mL of Tetramer and kept for 90 minutes at 37°C. Then, cells were washed twice with PBS and the wells were removed to reveal a microscope slide. The slide was fixed in acetone for 20 minutes at -20°C, followed by three washing steps with PBS. Next, the cells were blocked in 1x Carbo-free solution (Vector Laboratories) to prevent unspecific binding of the secondary antibody. After blocking, we stained with the primary unconjugated antibody anti-APC (clone eBioAPC-6A2, 1:100, ThermoFisher) for 1 hour at room temperature in the dark. We removed primary antibody, washed with PBS and then stained with goat anti-mouse AlexaFluor 647 (1:500, ThermoFisher) for 30 minutes at room temperature in the dark. The slide was washed again and then stained with anti-TCRβ conjugated with AlexaFluor 488 (clone H57-597, 1:100, BioLegend) for 1 hour at room temperature in the dark. We performed final washing steps and mounted the slide in Fluoroshield containing DAPI (Sigma Aldrich). The slide was kept at 4°C until further analysis. Images were acquired with a Leica Imager DMi8, while image analysis was performed with ImageJ software (Fiji).

### Immunophenotyping and tracking of ADRB1-specific T cells in PBMCs

PBMCs from DRB1*13^+^ patients recruited within the CAMI and ANTIK cohorts were thawed in a water bath at 37°C and washed twice with RPMI at 330 x *g* and room temperature for 5 minutes. Next, cells were subjected to magnetic activated cell sorting (MACS) for CD4^+^ T cells. In brief, cells in suspension were stained with Biotin-Antibody cocktail (biotin-conjugated antibodies against CD8a, CD14, CD15, CD16, CD19, CD36, CD56, CD123, TcRγ/δ and CD235a Miltenyi), followed by labelling with magnetic anti-biotin MicroBeads (Miltenyi). Subsequently, the cell suspension was loaded on MACS-LS columns and magnetic activated negative CD4^+^ T cell selection was performed according to the manufacturer’s instructions. The negatively selected CD4^+^ cells were resuspended in 200 μL of RPMI and plated in a U-bottom 96-well plate. After centrifugation, cells were resuspended in 50 μL of RPMI complete with 2 μg/mL of Tetramer and kept for 90 at 37°C. Then, cells were washed with RPMI and stained with efluor 780 viability dye for 15 minutes at room temperature. Next, cells were washed with FACS buffer and resuspended in 50 μL of antibody master mix, containing: anti-CD3 eFluor450 (clone UCHT1, ThermoFisher), - CD45 BUV496 (clone HI30, ThermoFisher), -CD4 Superbright 600 (clone SK3, ThermoFisher), -CCR7 (CD197) PerCP/Cy5.5 (clone G043H7, BioLegend), - CD45RA PE BUV737 (clone HI100, ThermoFisher), -CD25 FITC (clone M- A251, BioLegend), -CD127 PE-Cy7 (clone eBioRDR5, ThermoFisher), - CXCR3 BV480 (clone CXCR3-173, BD Biosciences), -CCR6 (CD196) BV650 (clone G034E3, BioLegend), -PD-1 PE-eFluor 610 (clone J105, ThermoFisher), -CD278 (ICOS) BUV395 (clone C398.4A /RUO, BD Biosciences), -CD69 PE-Fire640 (clone FN50, BioLegend), -CD134 (OX40) BV785 (clone Ber-ACT35/ACT35, BioLegend), -4-1BB PE (clone 4B4-1, ThermoFisher) and Human TruStain FcX™ (1:200, BioLegend).

Cells were kept for 30 minutes at 4°C and then washed with FACS buffer. Finally, the cells were resuspended in PBS and acquired on Cytek 5-Laser Aurora.

### Statistical analysis

Data are shown as mean ± SEM alongside the individual distribution of n datapoints. Statistical analysis was performed with an unpaired t-test, assuming normally distributed data for comparisons between two groups or One-way ANOVA with Dunnett’s multiple comparisons test for multiple groups. For comparisons within multiple groups with three subgroups each, statistical analysis used two-way ANOVA followed by Tukey’s multiple comparison test. Statistical analysis of allele frequency was performed with Chi-square test followed by Yates’ correction, and carrier frequency with Fisher’s exact test. *P*-values < 0.05 were considered statistically significant with: *, *P* <0.05; **, *P* < 0.01; ***, *P* < 0.001 and ****, *P* <0.0001. Statistical analysis was conducted using GraphPad Prism (version 10.2.1).

### Data accessibility

The anonymised patient data can be shared under reasonable request.

## Acknowledgments

We thank the NIH Tetramer Core Facility (NIH Contract 75N93020D00005 and RRID:SCR_026557) for providing the HLA-DRB1*13 tetramers used in this study. Some Figures were prepared using graphic elements freely available through the Servier Medical Art.

## Author contributions

Conceptualization: SF, GCR; Methodology: AR, MB, MDG, DA, TK, AES, KL, EMF; Investigation: AR, MB, MDG, DA, AH, MH, JS, TK, KL, EMF; Funding acquisition: SF, VN, GCR; Project administration: SF, GCR; Supervision: SF, GCR; Writing – original draft: AR, MDG, GCR; Writing – review & editing: all

## Extended Data Figures

**Extended Data Figure 1:**
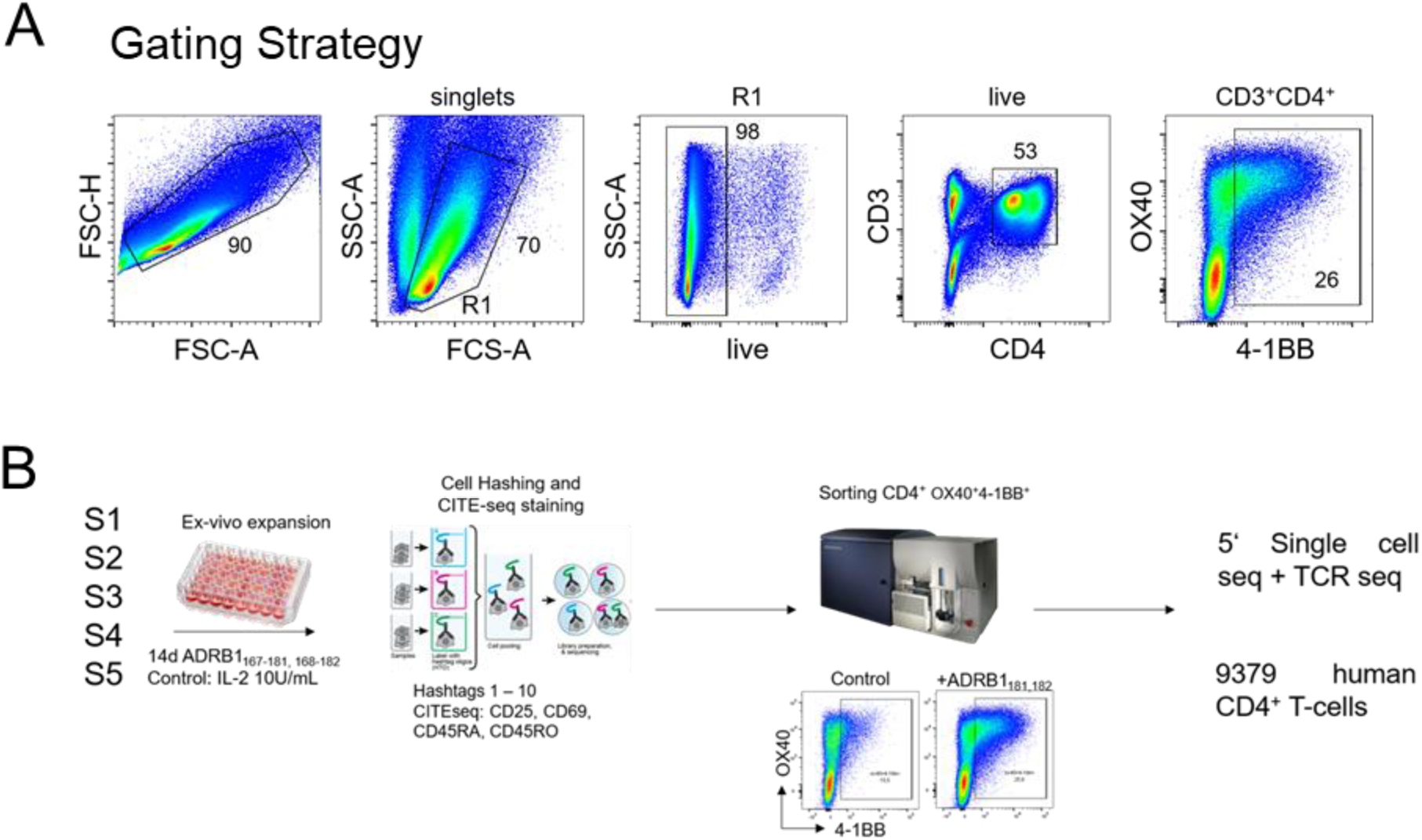
(**A**) Gating strategy and (**B**) sorting experiment design. PBMCs from five subjects (S1-S5) were kept for 14 days in culture in the presence of either IL-2 or IL-2 combined with ADRB1 peptides (10 conditions). Each condition was stained with a different hashtag antibody along with CITE-seq for CD69, CD25, CD45RA and CD45RO. Singlets, live, CD3^+^CD4^+^OX40^+/-^4-1BB^+^ events were sorted and cells from all conditions were pooled and sequenced as a single library. Following data filtration, 9,379 single cells were obtained.

**Extended Data Figure 2:**
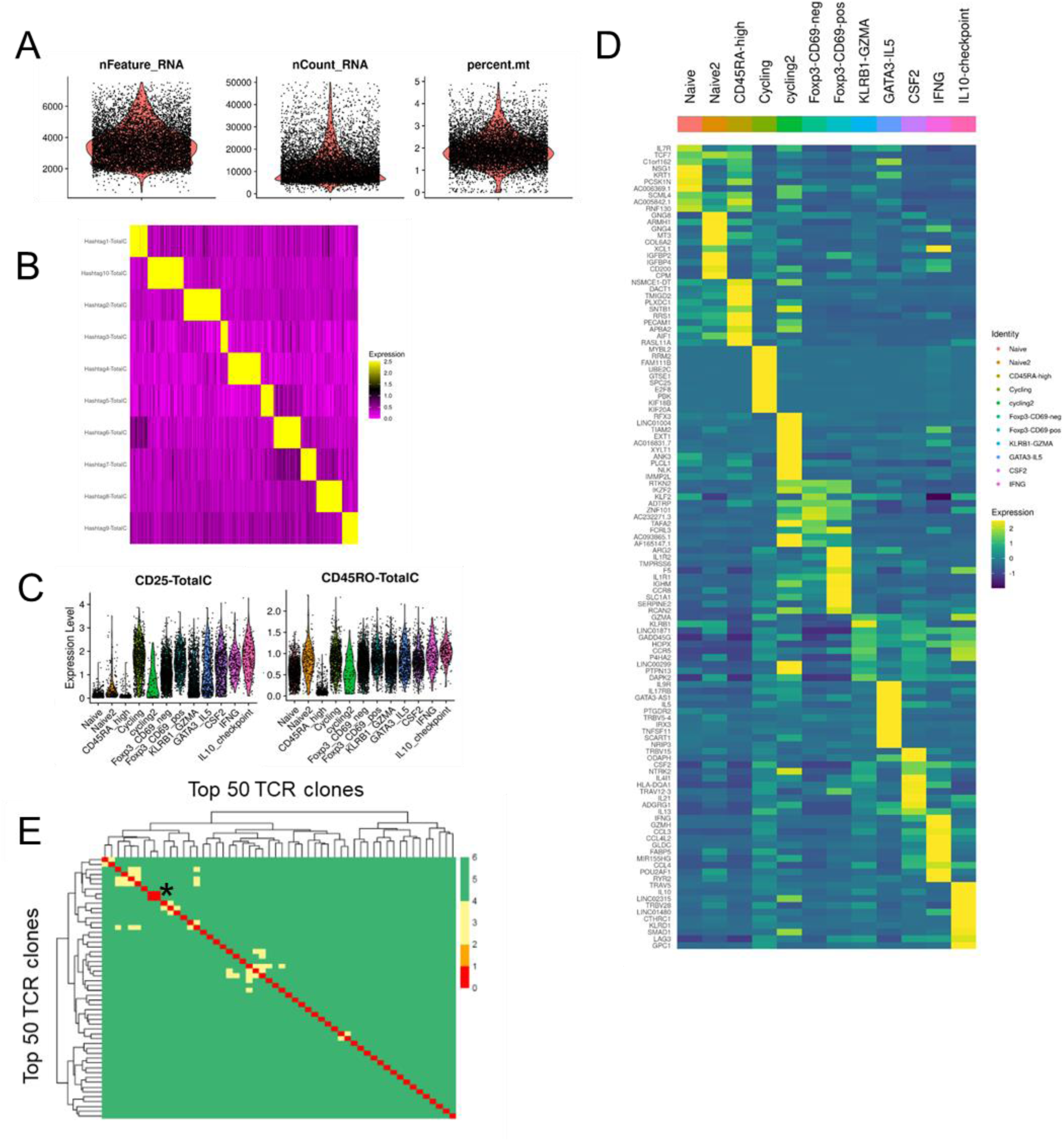
scRNA/ TCR sequencing quality control and top expressed transcripts per cluster. (**A**) Number of genes, counts and percentage of mitochondrial RNA detected in single cells. (**B**) Heatmap of hashtags 1-10, showing overall cell distributions. #1, 3, 5, 7, 9 show IL-2 control-stimulated cells from S1-S5, respectively, while #2, 4, 6, 8 and 19 show the ADRB1-stimulated cells from S1-S5. (**C**) Violin plots illustrate CITE-seq signal for CD25 and CD45RO. (**D**) Heatmap showing the top 10 up-regulated transcripts per cluster. (**E**) Heatmap shows the Levenshtein distance between the top 50 expanded CDR3β sequences derived from ADRB1-stimulated cultures. The * indicates the great similarity exhibited by the top two expanded clones found in S5.

**Extended Data Figure 3:**
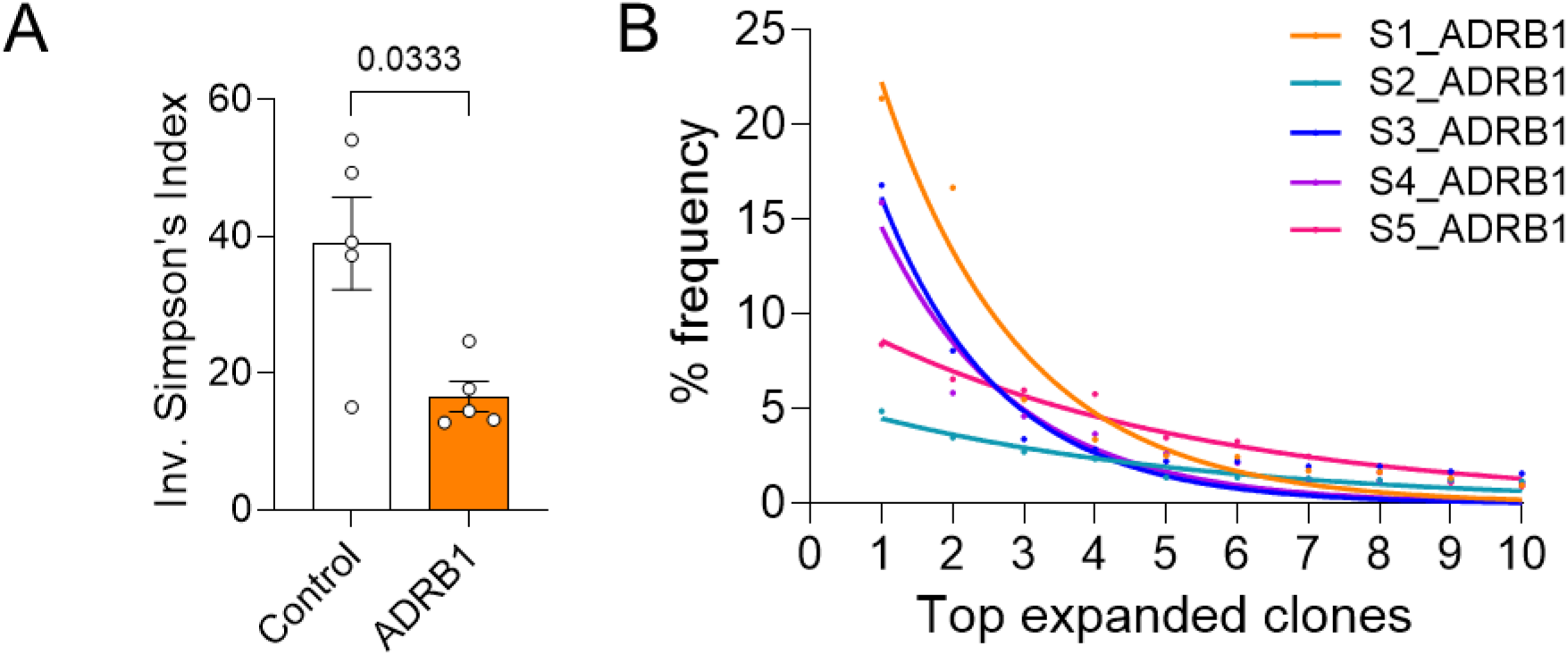
(**A**) Inverted Simpson’s index was estimated for each subject and plotted from control and peptide-stimulated group. (**B**) Frequency of top expanded clones in either ADRB1-stimulated group. Each line represents a different subject.

**Extended Data Figure 4:**
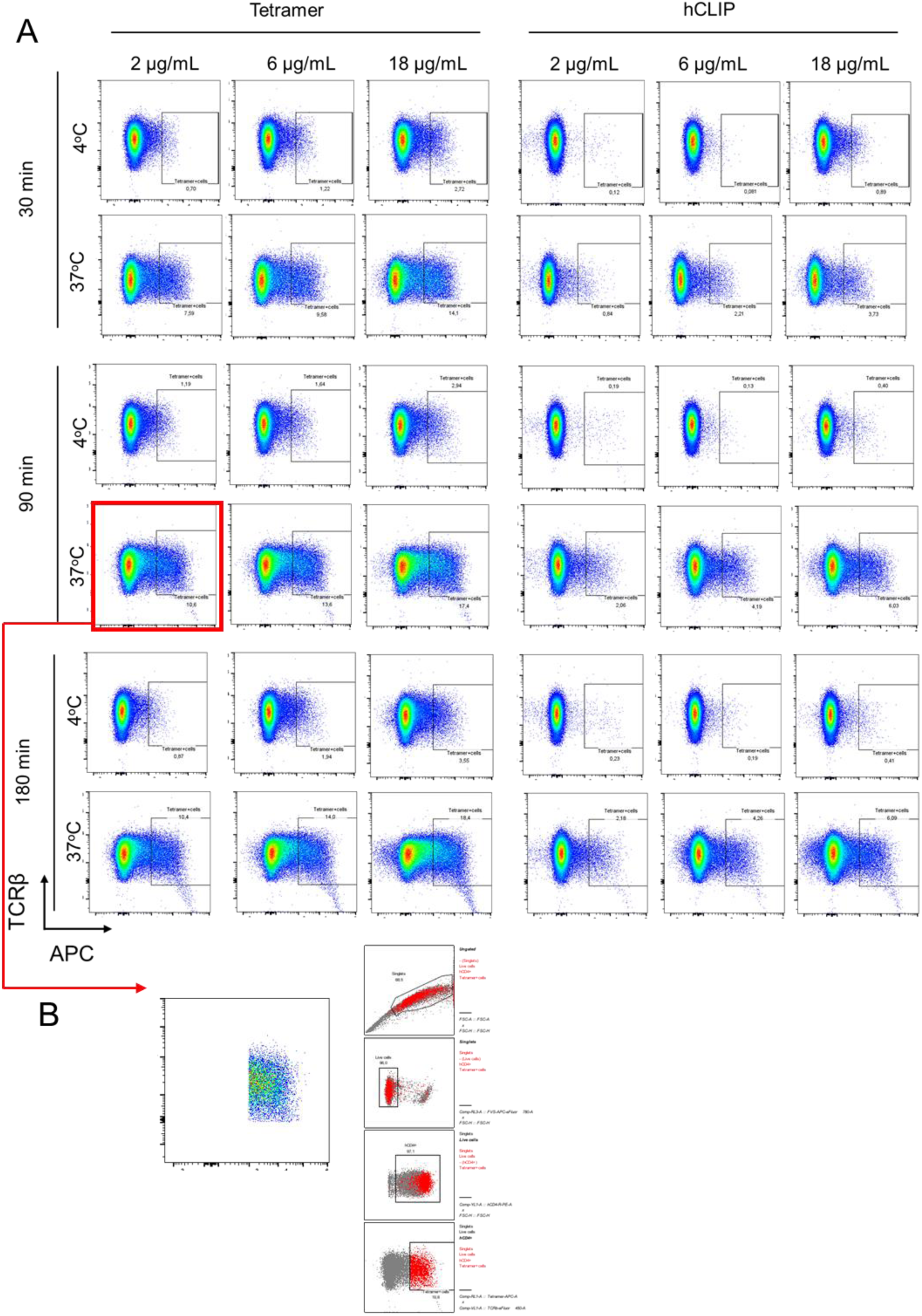
Establishing the experimental conditions of MHCII restricted tetramer against ADRB1_167-182_: (**A**) Representative flow cytometry plots of TCR6 cells (gated in live hCD4^+^) stained with three different concentrations (2, 6 and 18 μg/mL) of Tetramer or hCLIP for three different time points (30, 90 and 180 min) at two different temperatures (4 and 37 °C). The red square indicates the chosen experimental conditions. (**B**) Backgating of Tetramer^+^ cells.

**Extended Data Figure 5:**
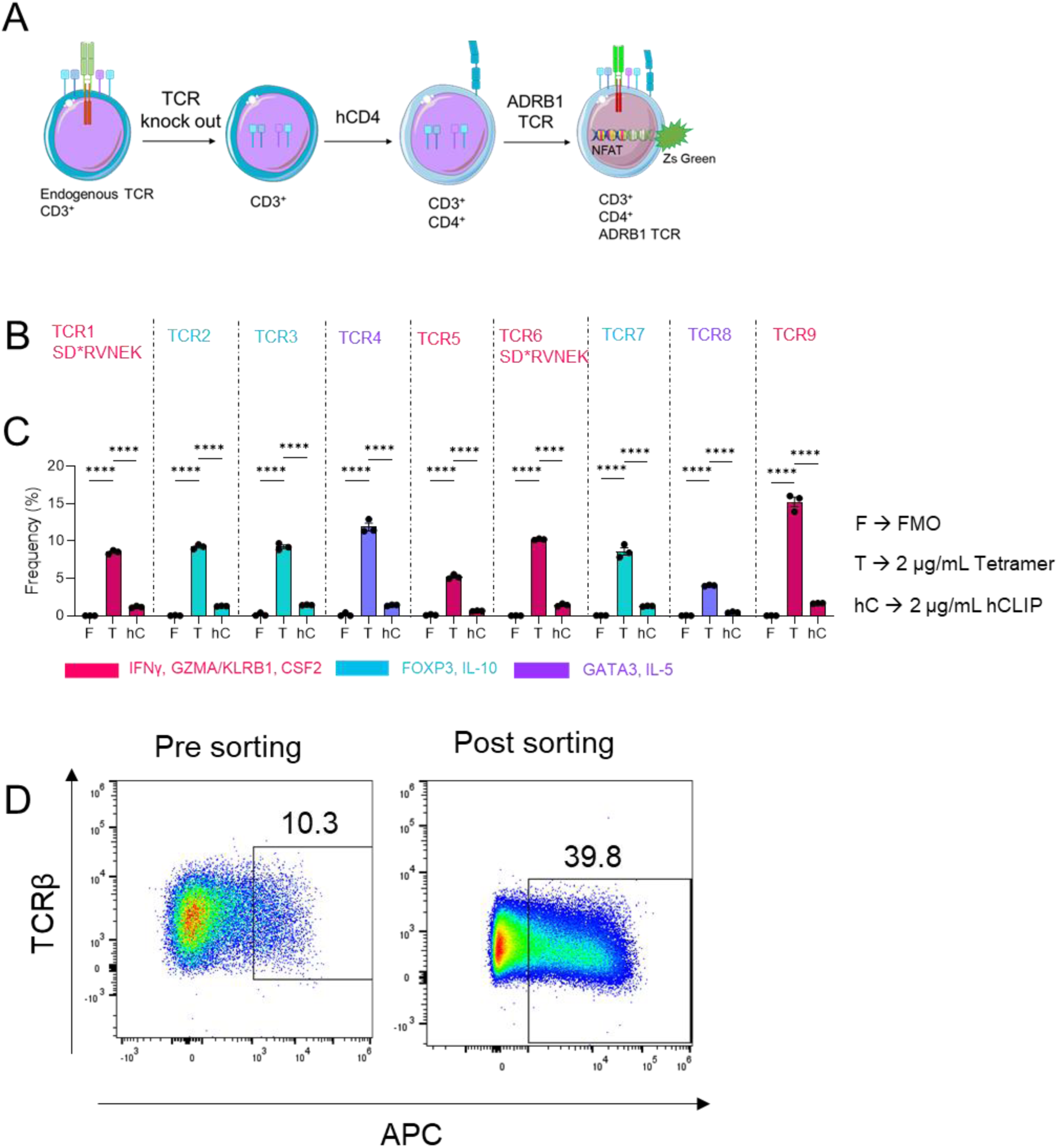
(**A**) Workflow of tgTCR cell line generation. (**B**) The nine generated tgTCRs coloured according to their corresponding cell cluster from scRNAseq (Figure 2B). TCR1 and TCR6 express the SD*RVNEK motif described in Figure 2. (**C**) Staining of tgTCRs with MHCII restricted tetramer against ADRB1_167-182_ and control hCLIP peptide. Unstained tgTCRs served as FMO (n=3 technical replicates; data are shown as mean±SEM; Statistical analysis was performed with two-way ANOVA followed by Tukey’s multiple comparison test. *P*-values < 0.05 were considered statistically significant with: ***, *P*<0.001; ****, *P*<0.0001). (**D**) Representative flow cytometry plots of TCR6 before sorting for tetramer^+^ cells (left) and after sorting (right).

**Extended Data Figure 6:**
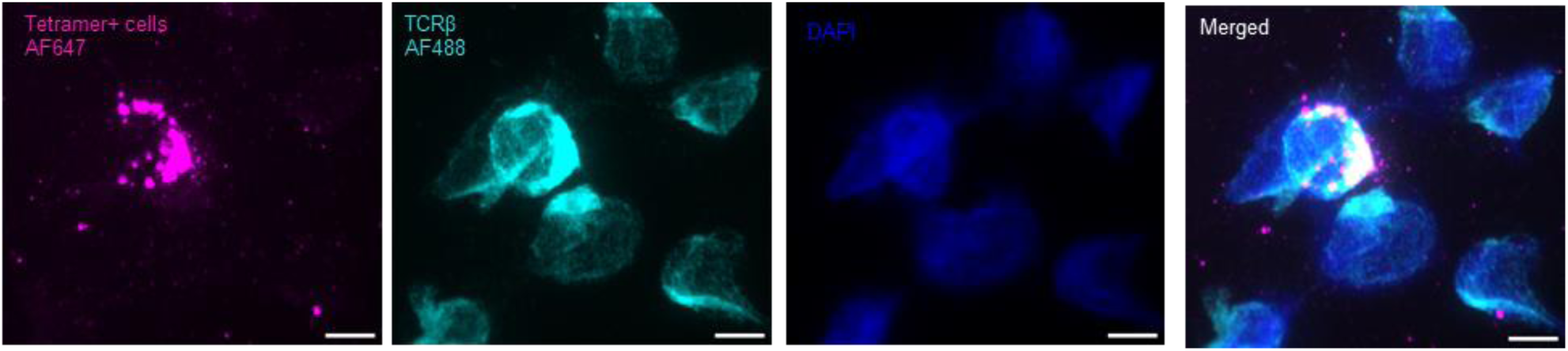
Representative fluorescent images of TCR6 cells stained by immunofluorescence for tetramer^+^ (magenta), TCRβ (cyan) and DAPI (blue). Images were acquired on a Leica Imager DMi8 equipped with a 63x objective. Scale bar: 10 μm.

**Extended Data Figure 7:**
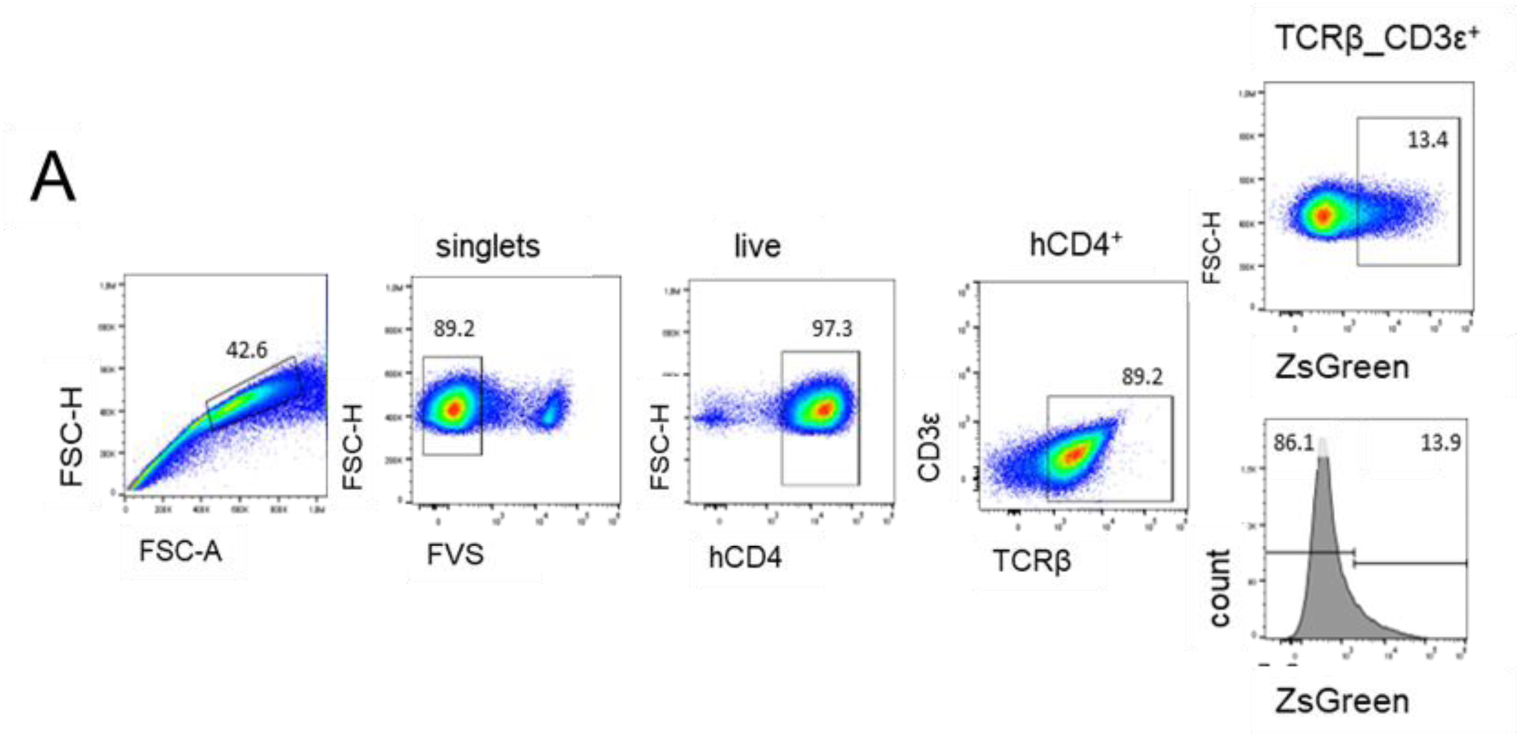
Gating strategy to analyse tgTCR activation after co-culture with PBMCs derived from DRB1*13^+^ subjects.

**Extended Data Figure 8:**
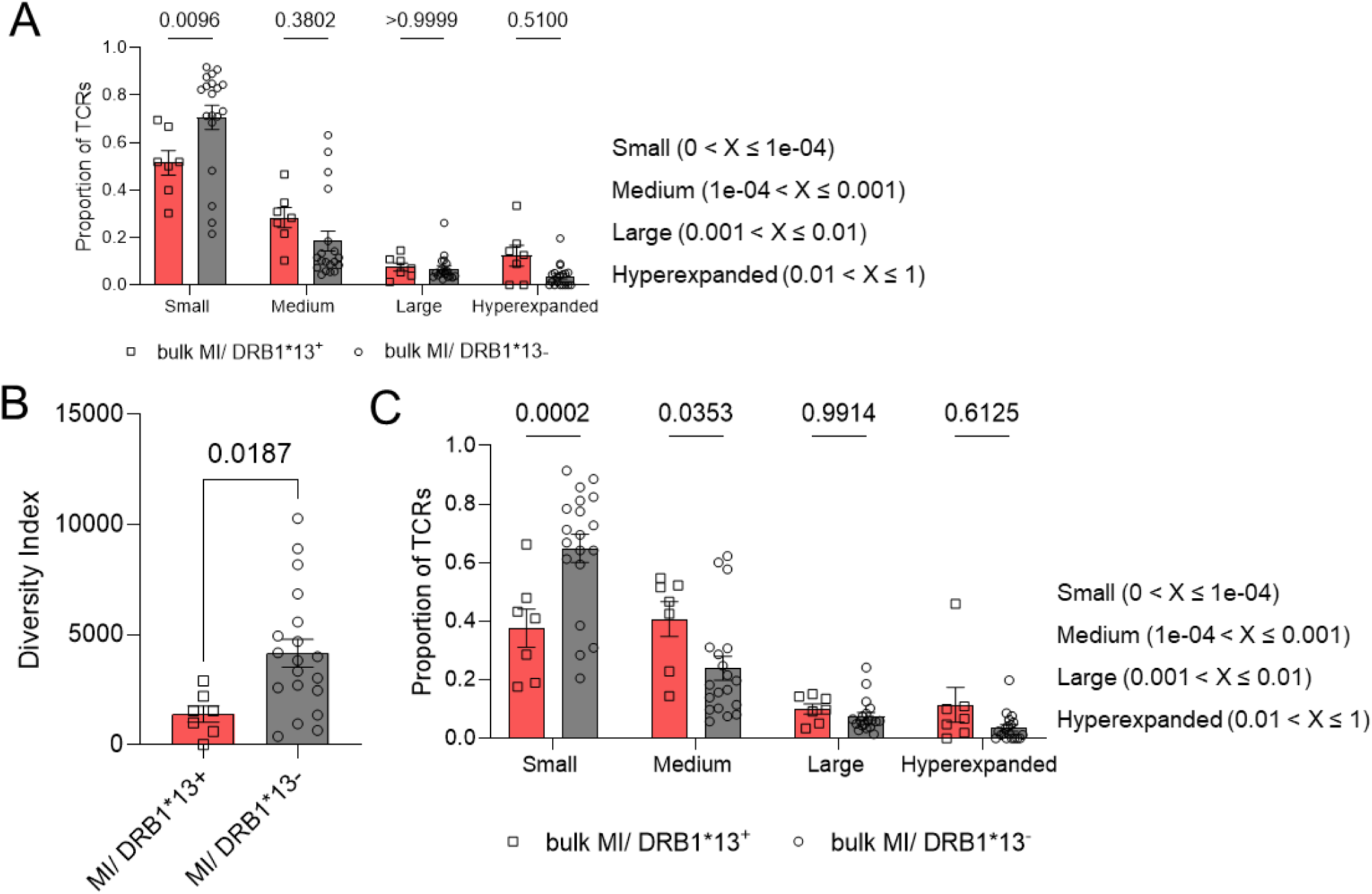
(**A**) Graph depicting expanded clone proportions in bulkRNAseq of DRB1*13^+^ (n=7) and DRB1*13^−^ (n=19) MI patients (Small: 0 < X ≤ 1e-04/, Medium: 1e-04 < X ≤ 0.001/, Large: 0.001 < X ≤ 0.01/, Hyperexpanded: 0.01 < X ≤ 1). Data are shown as mean±SEM; statistical analysis was performed with two-way ANOVA followed by Sidak’s multiple comparison test. *P*-values < 0.05 were considered statistically significant. (**B**) Diversity index of TCRα chain from the bulkRNAseq data of DRB1*13^+^ (n=7) versus DRB1*13^−^ (n=19) MI patients. Data are shown as mean±SEM; Statistical analysis was performed with unpaired t-test. *P*-values < 0.05 were considered statistically significant. (**C**) Graph depicting the proportion of expanded clones of α chain in bulkRNAseq of DRB1*13^+^ (n=7) and DRB1*13^−^ (n=21) MI patients (Small: 0 < X ≤ 1e-04/, Medium: 1e-04 < X ≤ 0.001/, Large: 0.001 < X ≤ 0.01/, Hyperexpanded: 0.01 < X ≤ 1). Data are shown as mean±SEM; statistical analysis was performed with two-way ANOVA followed by Sidak’s multiple comparison test. *P*-values < 0.05 were considered statistically significant.

**Extended Data Figure 9:**
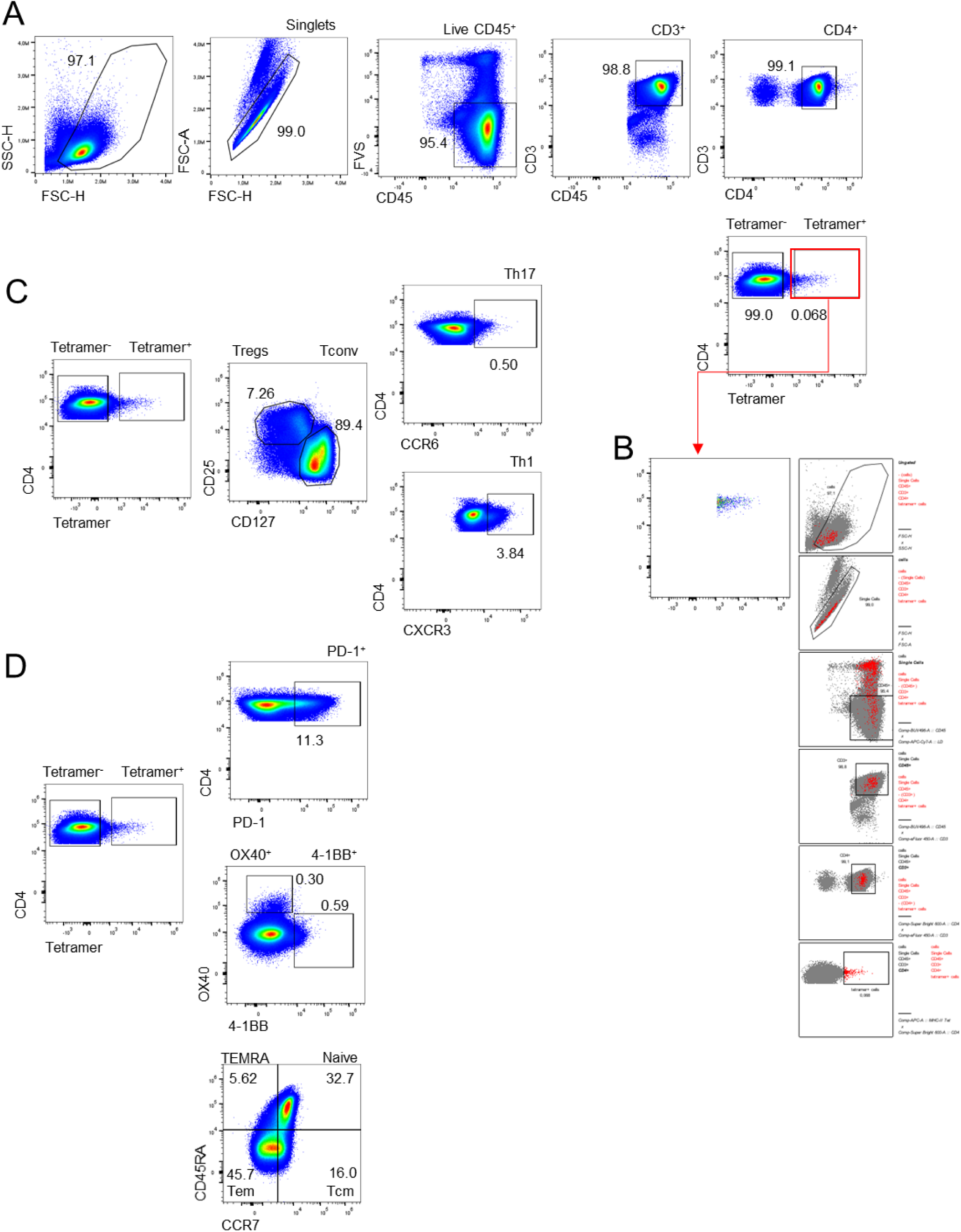
(**A**) Gating strategy to analyse tetramer^+^ and tetramer^−^ cells originated from MI DRB1*13^+^ patients. (**B**) Backgating of tetramer^+^CD4^+^ T cells. (**C**), (**D**) Gating strategy to further analyse tetramer^+^ and tetramer^−^ cells.

**Extended Data Figure 10:**
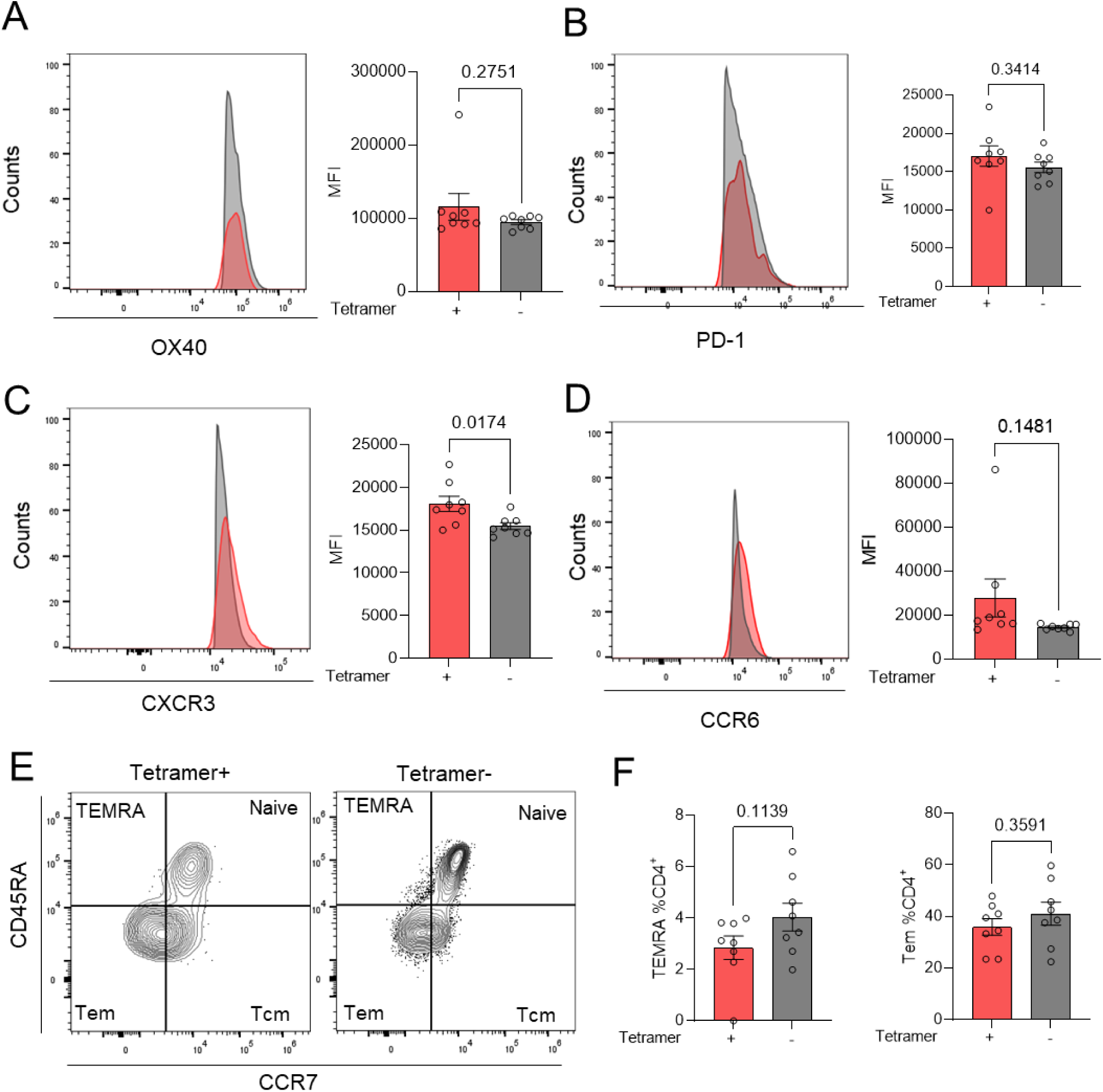
(**A**) Representative histogram (left) and quantitative analysis (right) of OX40^+^ tetramer^+^ versus tetramer^−^ cells for each patient (n=8). Data are shown as mean±SEM; Statistical analysis was performed with unpaired t-test. *P*-values < 0.05 were considered statistically significant. (**B**) Representative histogram (left) and quantitative analysis (right) of PD-1^+^ tetramer^+^ versus tetramer^−^ cells for each patient (n=8). Data are shown as mean±SEM; Statistical analysis was performed with unpaired t-test. *P*-values < 0.05 were considered statistically significant. (**C**) Representative histogram (left) and quantitative analysis (right) of Th1 (CXCR3^+^) Tetramer^+^ versus Tetramer^−^ cells for each patient (n=8). Data are shown as mean±SEM; Statistical analysis was performed with unpaired t-test. *P*-values < 0.05 were considered statistically significant. (**D**) Representative histogram (left) and quantitative analysis (right) of T_H_17 (CCR6^+^) tetramer^+^ versus tetramer^−^ cells for each patient (n=8). Data are shown as mean±SEM; Statistical analysis was performed with unpaired t-test. *P*-values < 0.05 were considered statistically significant. (**E**) Representative flow cytometry plot of CD45RA/CCR7 gated on live CD45^+^/CD3^+^/CD4^+^ cells and (**F**) quantitative analysis of T_EMRA_ and T_em_ cells in tetramer^+^ versus tetramer^−^ cells for each patient (n=8). Data are shown as mean±SEM; Statistical analysis was performed with unpaired t-test. *P*-values < 0.05 were considered statistically significant.

## Extended Data Tables

**Extended Data Table I:**
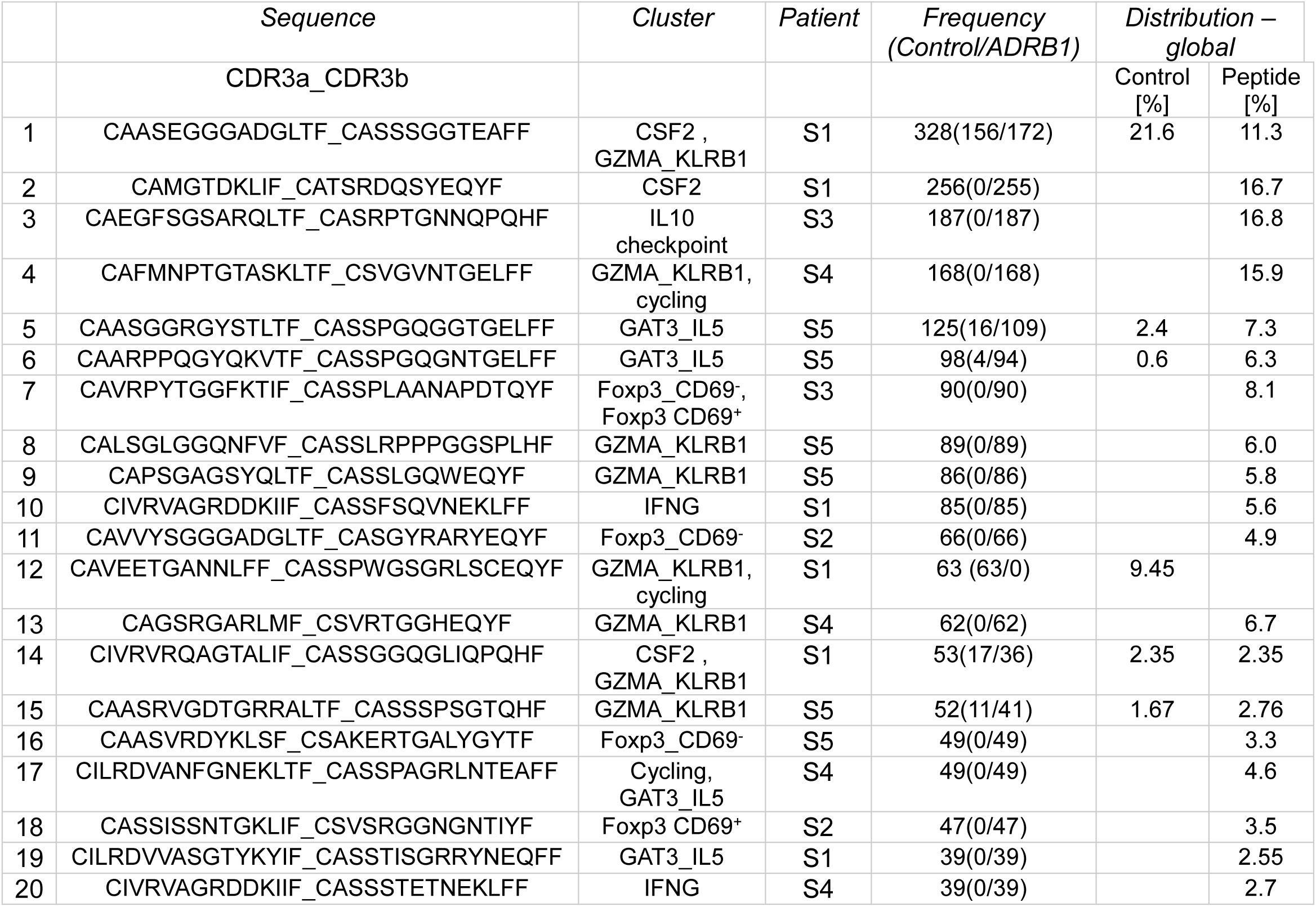
Top 20 TCR αβ sequences expanded in both control and ADRB1-stimulated PBMCs. For each sequence, the main representative cluster, subject, total counts and global distribution per subject and experiment condition are shown.

**Extended Data Table II:** Patient characteristics. CA coronary angiography, HS healthy subject.

| *Characteristics* |  | *CAMI* |  |  | *ETiCS* | *ANTIK* |
| --- | --- | --- | --- | --- | --- | --- |
|  |  | *MI (n=7)* | *CA (n= 2)* | *HS*<br>*(n=1)* | *MI (n=26)* | *MI (n=2)* |
| *sex* | male | 6 (0.86) | 2 (1) | 0 (0) | 30 (0.97) | 2 (1) |
|  | female | 1 (0.14) | 0(0) | 1 (1) | 1 (0.03) | 0 (0) |
| *Haplotype* | HLADRB1*-<br>13<sup>+</sup> | 7<br>(1.00) | 2<br>(1.00) | 1<br>(1.00) | 7<br>(0.27) | 2<br>(1) |
|  | HLADRB1*-<br>13<sup>-</sup> | 0<br>(0.00) | 0<br>(0.00) | 0<br>(0.00) | 19<br>(0.73) | 0<br>(0) |
| *type of MI* | STEMI | 6 (0.86) | n.a. | n.a. | 26(1) | 2 (1) |
|  | NSTEMI | 1 (0.14) | n.a. | n.a. | 0(0) | 0 |
| *1st troponin*<br>*(pg/ml)* | mean +/-<br>SEM | 572.51<br>+/- 175.64 | n.a. | n.a. | 2,647.17<br>+/- 288.60 | 3,187.9<br>+/- 2,226.75 |
| *Type of*<br>*biomaterial* |  | PBMCs |  |  | whole blood -<br>DMSO | PBMCs |
| *Time of*<br>*sampling* |  | day 2-8 | n.a. |  | index<br>hospitalization | day 5 |

